# Adverse Events Following SARS-CoV-2 mRNA Vaccination in Adolescents: A Norwegian Nationwide Register-Based Study

**DOI:** 10.1101/2023.12.13.23299926

**Authors:** Vilde Bergstad Larsen, Nina Gunnes, Jon Michael Gran, Jesper Dahl, Håkon Bøås, Sara Viksmoen Watle, Jacob Dag Berild, Margrethe Greve-Isdahl, Ketil Størdal, Hanne Løvdal Gulseth, Øystein Karlstad, Paz Lopez-Doriga Ruiz, German Tapia

## Abstract

**Background:** Vaccination of older adolescents against severe acute respiratory syndrome coronavirus 2 (SARS-CoV-2) started in the spring of 2021 and continued with younger adolescents throughout the summer and fall. We assessed risks of adverse events following immunization (AEFI) in adolescents aged 12–19 years following SARS-CoV-2 vaccination with a messenger RNA (mRNA) vaccine in Norway.

**Materials and Methods:** The study sample included 496,432 adolescents born in 2002–2009, residing in Norway, and unvaccinated against SARS-CoV-2 at the beginning of the age-specific waves of vaccination in 2021. The exposures under study were first- and second-dose SARS-CoV-2 mRNA vaccinations vs. no dose. We applied Poisson regression and self-controlled case series (SCCS) analysis to estimate incidence rate ratios (IRRs) of 17 preselected outcomes, with associated 95% confidence intervals (CIs), between vaccinated and unvaccinated subjects using predefined post-vaccination risk windows.

**Results:** Most outcome-specific numbers of cases were low. There were no statistically significant associations between first-dose vaccination and any of the outcomes. In the main Poisson regression, second-dose vaccination was associated with increased risks of anaphylactic reaction (adjusted IRR [aIRR]: 10.05; 95% CI: 1.22–82.74), lymphadenopathy (aIRR: 2.33; 95% CI: 1.46–3.72), and myocarditis and pericarditis (aIRR: 5.27; 95% CI: 1.98–14.05). We also observed increased incidence of acute appendicitis outside the 14-day risk window. When expanding the risk window to 42 days in a post-hoc analysis, there was increased incidence of acute appendicitis following both first-dose vaccination (aIRR: 1.39; 95% CI: 1.09–1.78) and second-dose vaccination (aIRR: 1.43; 95% CI: 1.07–1.91). Results of the SCCS analysis were similar to the Poisson regression.

**Conclusions:** In general, potential AEFI were rare among adolescents. We found increased risks of anaphylactic reaction, lymphadenopathy, and myocarditis and pericarditis following second-dose vaccination. There were also indications of increased acute appendicitis risk when applying longer risk windows.

## Introduction

Although severe acute respiratory syndrome coronavirus 2 (SARS-CoV-2) infection is milder in adolescents,[1] severe disease and postinfectious conditions may develop.[2, 3] The Norwegian COVID-19 vaccination campaign offered healthy adolescents born in 2002–2009 vaccination from April 2021, while those with chronic conditions were vaccinated prior. The two messenger RNA (mRNA) vaccines Comirnaty (BNT162b2, Pfizer-BioNTech) and Spikevax (mRNA-1273, Moderna) were widely administered in Norway, with adolescents predominantly receiving Comirnaty.

Phase 3 clinical trials of mRNA vaccines in adolescents and adults reported increased risks of lymphadenopathy (16+ years) and Bell’s palsy (18+ years),[4–6] and the proportions of appendicitis, acute myocardial infarction, and cerebrovascular accidents were higher among adults.[4] Vaccine trials of Comirnaty in children (5–11 years, *n*=2,268) and adolescents (12–15 years, *n*=2,260) and of Spikevax in adolescents (12–17 years, *n*=3,732) did not report any vaccine-related serious adverse events.[7–9]

Clinical trials are by necessity limited in size and tend to include healthier subjects, which might not reflect real-world settings. Large cohort studies are therefore essential for post-marketing safety surveillance of potential adverse events. A large Israeli population-based study (16+ years) found the Comirnaty vaccine to be associated with myocarditis, lymphadenopathy, appendicitis, and herpes zoster.[10] A recent systematic review on adverse events after SARS-CoV-2 vaccination in adolescents reported myocarditis and pericarditis as most concerning.[11] Similar incidence rates were reported for acute allergic reaction and convulsions, although in fewer studies.[11] Further investigations of potential adverse events following SARS-CoV-2 mRNA vaccination in adolescents are warranted to fill knowledge gaps and ensure trust in the vaccines.

The aim of this nationwide study was to assess short- and mid-term safety of SARS-CoV-2 mRNA vaccines in all adolescents aged 12–19 years in Norway by exploring associations with preselected adverse events.

## Materials and Methods

This study was based on the Norwegian Emergency Preparedness Register for COVID-19 (Beredt C19).[12] Beredt C19 includes individual-level data on demographics, SARS-CoV-2 infections and vaccinations, and diagnoses from primary and specialist health services, linkable by the unique national identity number assigned to all residents. For further details, see the Supplementary Materials.

### Outcomes

Experienced clinicians within our research group identified 17 outcomes of interest following SARS-CoV-2 vaccination based on mRNA vaccine trials, Norwegian COVID-19 surveillance, and adverse events reported by healthcare professionals. Outcome-specific risk windows were based on recommendations from the World Health Organization, European Medicines Agency, and Brighton Collaboration. Table 1 lists the outcomes with corresponding diagnosis codes and risk windows.

**Table 1:** Conditions and the corresponding ICD-10^1^ codes that define the outcomes of interest.

| Condition | ICD-10<sup>1</sup> codes | Risk window | Inpatient/outpatient |
| --- | --- | --- | --- |
| Acute appendicitis | K35 | 14 days | Inpatient |
| Anaphylactic reaction | R57.9, T78.2, T88.6 | 2 days | Inpatient |
| Arrhythmia | I44, I45, I46, I47, I48, I49 | 28 days | Inpatient |
| Arthropathy | M02.2, M02.9 | 42 days | In- and outpatient |
| Cerebrovascular events, combined | G45, I60, I61, I62, I63, I64 | 28 days | Inpatient |
| Death<sup>2</sup> | - | 28 days | - |
| Encephalomyelitis and meningitis, combined | G03.0, G04.0, G04.8, G04.9, G36.9 | 28 days | Inpatient |
| Epilepsy and convulsions, combined | G40, G41, R56 | 28 days | In- and outpatient |
| Facial nerve palsy | G51.0, G51.9 | 28 days | In- and outpatient |
| Guillain-Barré syndrome | G61.0 | 42 days | Inpatient |
| Henoch-Schönlein purpura | D69.0, M36.4, N08.2 | 42 days | Inpatient |
| Herpes zoster | B02, S70<sup>3</sup> | 28 days | In- and outpatient |
| Idiopathic thrombocytopenic purpura | D69.3 | 28 days | In- and outpatient |
| Lymphadenopathy | I88.0, I88.8, I88.9, L04, R59 | 14 days | In- and outpatient |
| Multisystem inflammatory syndrome in children | B94.8, M35.8, U10.9 | 42 days | Inpatient |
| Myocarditis and pericarditis, combined | I30.0, I30.8, I30.9, I40.1, I40.8, I40.9, I51.4 | 28 days | Inpatient |
| Venous thromboembolic events, combined | I26, I63.6, I67.6, I80.1, I80.2, I80.3, I81, I82.0, I82.2, I82.3, I82.8, I82.9 | 28 days | In- and outpatient |

<sup>1</sup>International Statistical Classification of Diseases and Related Health Problems, Tenth Revision
<sup>2</sup>Date of death obtained from the Norwegian Population Register
<sup>3</sup>Code based on the International Classification of Primary Care, Second Edition (ICPC-2)

### Exposures

Exposures were first- and second-dose SARS-CoV-2 mRNA vaccinations (Comirnaty or Spikevax) vs. unvaccinated. National vaccination recommendations differed across age groups; 12–15-year-olds (born 2006–2009) were offered the first dose September 2021 and the second dose January 2022, whereas 16–17-year-olds (born 2004–2005) and 18–19-year-olds (born 2002–2003) were recommended two doses with an interval of 8–12 and 6–12 weeks, respectively.

### Study Sample

The study sample included 496,432 adolescents born in 2002–2009, residing in Norway (since January 1, 2017, or earlier) and unvaccinated against SARS-CoV-2 when the age-specific vaccination waves started: September 6, 2021 (12–15 years), August 23, 2021 (16–17 years), and April 5, 2021 (18–19 years).

### Statistical Analysis

We used two different methodological approaches to compare outcome-specific incidence rates between vaccinated and unvaccinated subjects: 1) cohort analysis with Poisson regression and 2) self-controlled case series (SCCS) analysis. Follow-up started on the first day of the respective vaccination wave of the three aforementioned age groups (see Study Sample). In each separate analysis, we used a washout period from January 1, 2017, excluding all subjects with the outcome in question the last four years prior to start of follow-up. Censoring events were non-mRNA vaccination, third-dose vaccination, emigration, death, or study end on September 30, 2022.

We defined a time-varying five-level categorical exposure variable for the subjects’ current mRNA vaccination status: 1) unvaccinated (reference group), 2) vaccinated with *first* dose and *inside* the risk window, 3) vaccinated with *first* dose and *outside* the risk window, 4) vaccinated with *second* dose and *inside* the risk window, and 5) vaccinated with *second* dose and *outside* the risk window. Subjects receiving a second dose while still inside the risk window following the first dose were classified as category 4. Our primary interest was outcomes inside the risk windows following vaccinations (categories 2 and 4, respectively).

### Poisson Regression

In the main analysis, we applied Poisson regression to estimate incidence rate ratios (IRRs) of each outcome, with associated 95% confidence intervals (CIs), for subjects after first- and second-dose vaccinations compared to non-vaccinees. Subjects were followed until the outcome in question or censoring, whichever occurred first. We adjusted for sex (male or female), age group (12–15, 16–17, or 18–19 years), health region (North, Central, West, or South-East Norway), and risk group (no vs any) based on preexisting conditions.[1] Since infection rates varies over time and outcomes may exhibit seasonality, we also adjusted for a time-varying categorical variable for the current calendar period (April–June 2021, July–September 2021, October–December 2021, January–March 2022, April–June 2022, and July–September 2022).

To assess associations in previously uninfected subjects, we conducted a sensitivity analysis where SARS-CoV-2 infection was a censoring event. We also conducted subgroup analyses based on attained age by end of 2021 to explore a potential effect modification by age group.

### SCCS Analysis

For each outcome, we also conducted an SCCS analysis of all incident cases to estimate IRRs, with associated 95% CIs, for time periods following first- and second-dose vaccinations compared to the unexposed control period, i.e., follow-up time before first-dose vaccination (Figure 1), as described by Whitaker et al.[13, 14] The cases were followed until censoring. We adjusted for seasonality (January–March, April–June, July–September, or October–December). No adjustment was made for age, as it in practice was considered time-invariant due to the relatively short study period.

**Figure 1:**
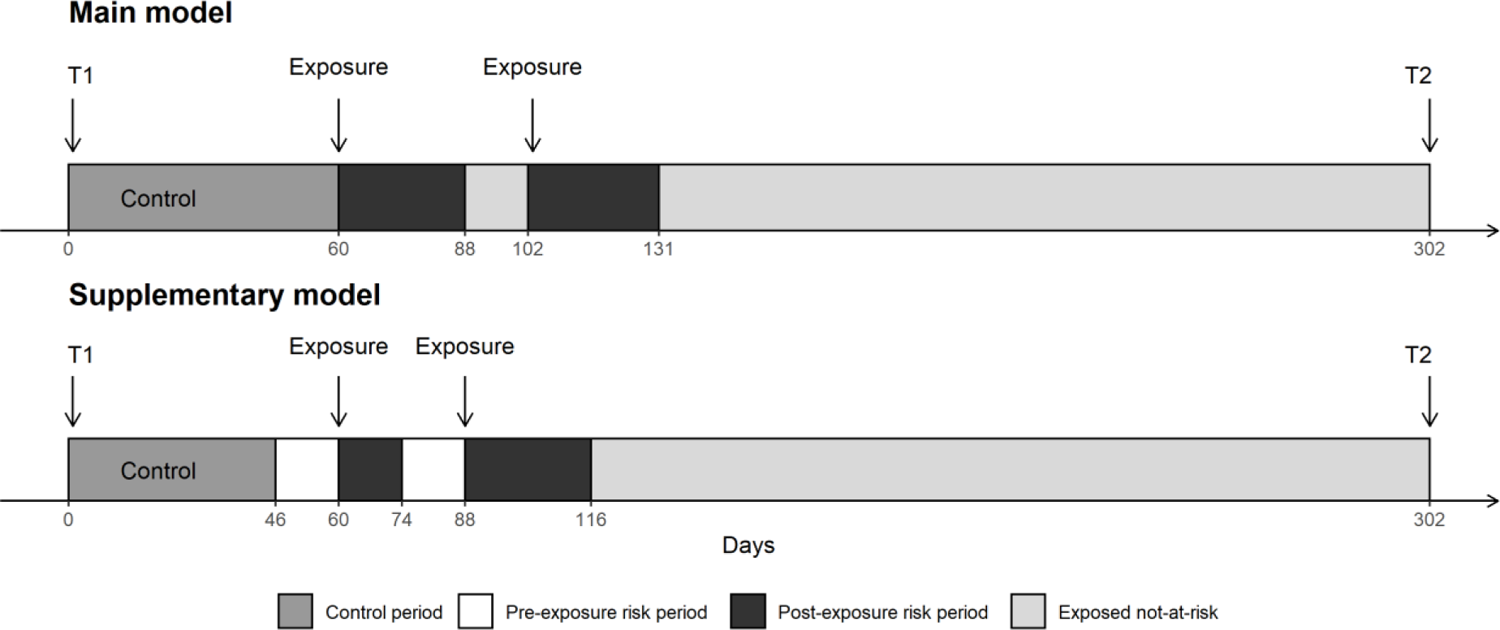
Hypothetical follow-up period in the self-controlled case series analysis based on the main model (upper panel) and the supplementary model with a 14-day preexposure risk period (lower panel). The postexposure risk period is defined by the outcome-specific risk windows.

We also conducted outcome-specific sensitivity analyses where we included 14-day pre-vaccination risk periods.[13] We then visually inspected exposure-centered plots to assess potential violation of the key assumption of outcomes not affecting future vaccination. The 14-day time period was considered sufficient to capture a potential healthy-vaccinee effect.

## Results

Table 2 displays cohort characteristics. The proportions of 12–15-year-olds, 16–17-year-olds, and 18–19-year-olds were 51.1%, 24.4%, and 24.5%, respectively, with a slight male predominance (51.4%). Preexisting risk conditions were found in 8.3%.

**Table 2:**
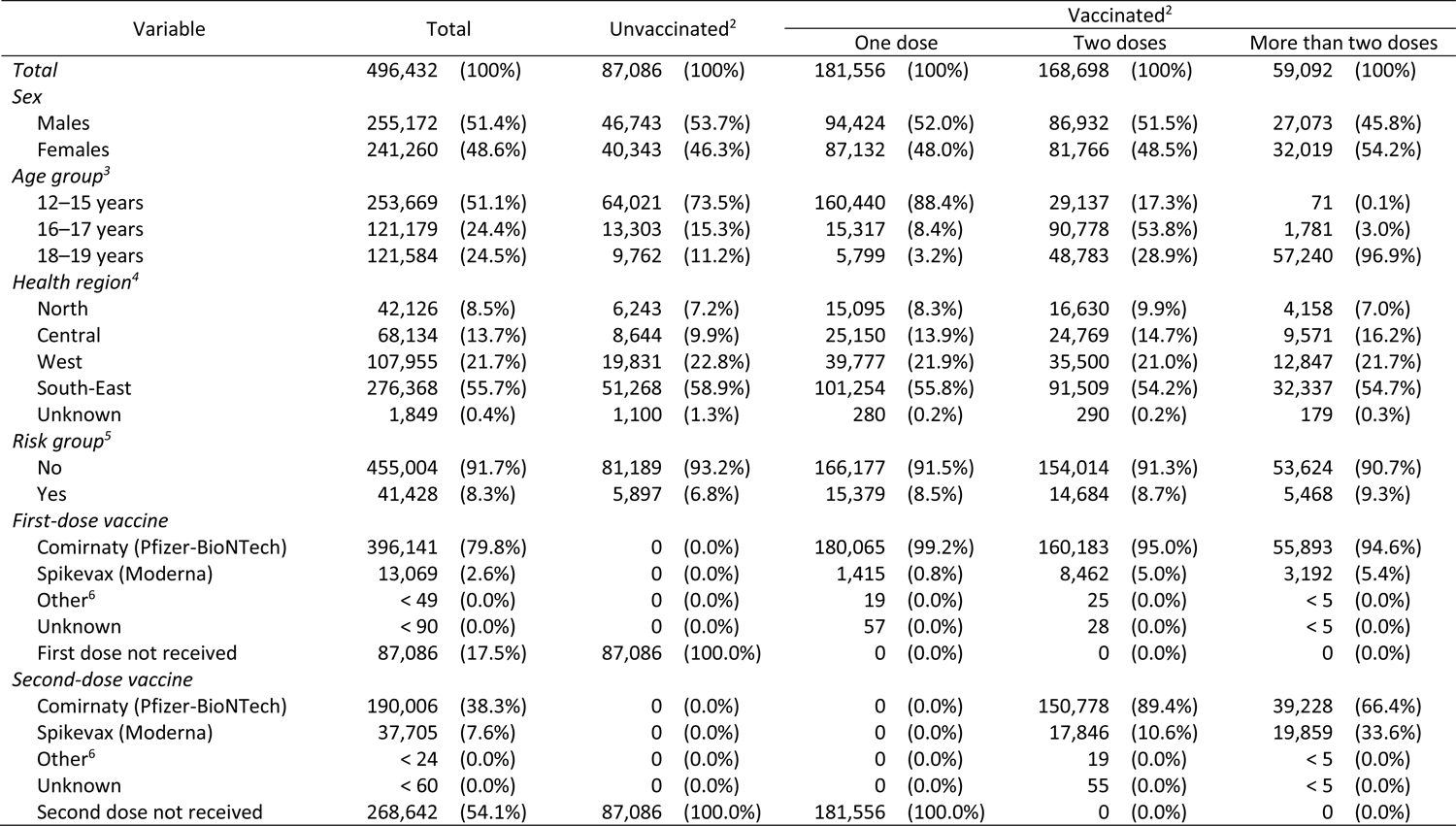
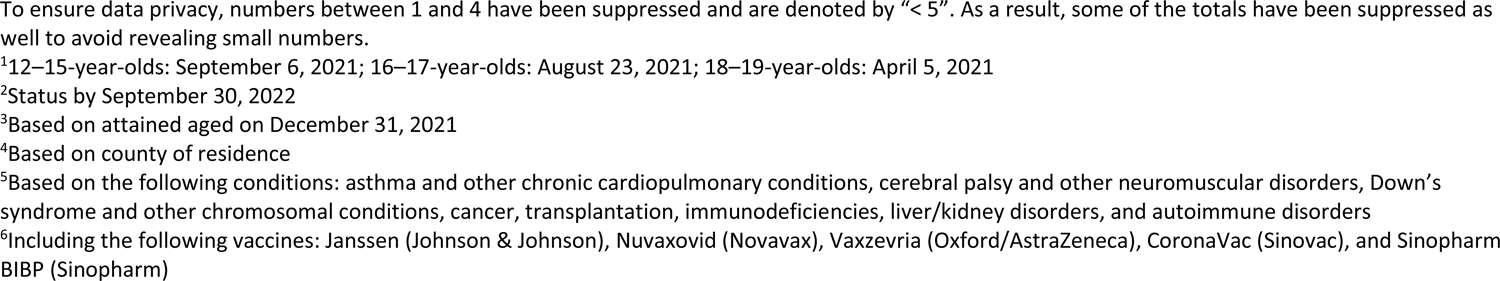
Characteristics of 496,432 adolescents born in 2002–2009 and still alive, residing in Norway (since January 1, 2017, or earlier), and unvaccinated against SARS-CoV-2 at the beginning of the wave of vaccination^1^ of their age group.

| Variable | Total |  | Unvaccinated<sup>2</sup> |  | Vaccinated<sup>2</sup> |  |  |  |  |  |
| --- | --- | --- | --- | --- | --- | --- | --- | --- | --- | --- |
|  |  |  |  |  | One dose |  | Two doses |  | More than two doses |  |
| *Total* | 496,432 | (100%) | 87,086 | (100%) | 181,556 | (100%) | 168,698 | (100%) | 59,092 | (100%) |
| *Sex* |  |  |  |  |  |  |  |  |  |  |
| Males | 255,172 | (51.4%) | 46,743 | (53.7%) | 94,424 | (52.0%) | 86,932 | (51.5%) | 27,073 | (45.8%) |
| Females | 241,260 | (48.6%) | 40,343 | (46.3%) | 87,132 | (48.0%) | 81,766 | (48.5%) | 32,019 | (54.2%) |
| *Age group<sup>3</sup>* |  |  |  |  |  |  |  |  |  |  |
| 12–15 years | 253,669 | (51.1%) | 64,021 | (73.5%) | 160,440 | (88.4%) | 29,137 | (17.3%) | 71 | (0.1%) |
| 16–17 years | 121,179 | (24.4%) | 13,303 | (15.3%) | 15,317 | (8.4%) | 90,778 | (53.8%) | 1,781 | (3.0%) |
| 18–19 years | 121,584 | (24.5%) | 9,762 | (11.2%) | 5,799 | (3.2%) | 48,783 | (28.9%) | 57,240 | (96.9%) |
| *Health region<sup>4</sup>* |  |  |  |  |  |  |  |  |  |  |
| North | 42,126 | (8.5%) | 6,243 | (7.2%) | 15,095 | (8.3%) | 16,630 | (9.9%) | 4,158 | (7.0%) |
| Central | 68,134 | (13.7%) | 8,644 | (9.9%) | 25,150 | (13.9%) | 24,769 | (14.7%) | 9,571 | (16.2%) |
| West | 107,955 | (21.7%) | 19,831 | (22.8%) | 39,777 | (21.9%) | 35,500 | (21.0%) | 12,847 | (21.7%) |
| South-East | 276,368 | (55.7%) | 51,268 | (58.9%) | 101,254 | (55.8%) | 91,509 | (54.2%) | 32,337 | (54.7%) |
| Unknown | 1,849 | (0.4%) | 1,100 | (1.3%) | 280 | (0.2%) | 290 | (0.2%) | 179 | (0.3%) |
| *Risk group<sup>5</sup>* |  |  |  |  |  |  |  |  |  |  |
| No | 455,004 | (91.7%) | 81,189 | (93.2%) | 166,177 | (91.5%) | 154,014 | (91.3%) | 53,624 | (90.7%) |
| Yes | 41,428 | (8.3%) | 5,897 | (6.8%) | 15,379 | (8.5%) | 14,684 | (8.7%) | 5,468 | (9.3%) |
| *First-dose vaccine* |  |  |  |  |  |  |  |  |  |  |
| Comirnaty (Pfizer-BioNTech) | 396,141 | (79.8%) | 0 | (0.0%) | 180,065 | (99.2%) | 160,183 | (95.0%) | 55,893 | (94.6%) |
| Spikevax (Moderna) | 13,069 | (2.6%) | 0 | (0.0%) | 1,415 | (0.8%) | 8,462 | (5.0%) | 3,192 | (5.4%) |
| Other<sup>6</sup> | < 49 | (0.0%) | 0 | (0.0%) | 19 | (0.0%) | 25 | (0.0%) | < 5 | (0.0%) |
| Unknown | < 90 | (0.0%) | 0 | (0.0%) | 57 | (0.0%) | 28 | (0.0%) | < 5 | (0.0%) |
| First dose not received | 87,086 | (17.5%) | 87,086 | (100.0%) | 0 | (0.0%) | 0 | (0.0%) | 0 | (0.0%) |
| *Second-dose vaccine* |  |  |  |  |  |  |  |  |  |  |
| Comirnaty (Pfizer-BioNTech) | 190,006 | (38.3%) | 0 | (0.0%) | 0 | (0.0%) | 150,778 | (89.4%) | 39,228 | (66.4%) |
| Spikevax (Moderna) | 37,705 | (7.6%) | 0 | (0.0%) | 0 | (0.0%) | 17,846 | (10.6%) | 19,859 | (33.6%) |
| Other<sup>6</sup> | < 24 | (0.0%) | 0 | (0.0%) | 0 | (0.0%) | 19 | (0.0%) | < 5 | (0.0%) |
| Unknown | < 60 | (0.0%) | 0 | (0.0%) | 0 | (0.0%) | 55 | (0.0%) | < 5 | (0.0%) |
| Second dose not received | 268,642 | (54.1%) | 87,086 | (100.0%) | 181,556 | (100.0%) | 0 | (0.0%) | 0 | (0.0%) |

To ensure data privacy, numbers between 1 and 4 have been suppressed and are denoted by “< 5”. As a result, some of the totals have been suppressed as well to avoid revealing small numbers.
<sup>1</sup>12–15-year-olds: September 6, 2021; 16–17-year-olds: August 23, 2021; 18–19-year-olds: April 5, 2021
<sup>2</sup>Status by September 30, 2022
<sup>3</sup>Based on attained aged on December 31, 2021
<sup>4</sup>Based on county of residence
<sup>5</sup>Based on the following conditions: asthma and other chronic cardiopulmonary conditions, cerebral palsy and other neuromuscular disorders, Down’s syndrome and other chromosomal conditions, cancer, transplantation, immunodeficiencies, liver/kidney disorders, and autoimmune disorders
<sup>6</sup>Including the following vaccines: Janssen (Johnson & Johnson), Nuvaxovid (Novavax), Vaxzevria (Oxford/AstraZeneca), CoronaVac (Sinovac), and Sinopharm BIBP (Sinopharm)

Figure 2 displays age-specific Kaplan-Meier estimates of the cumulative proportions of first- and second-dose SARS-CoV-2 mRNA vaccinations plotted against calendar time. There was a rapid increase in vaccination coverage the first months of the vaccination waves.

**Figure 2:**
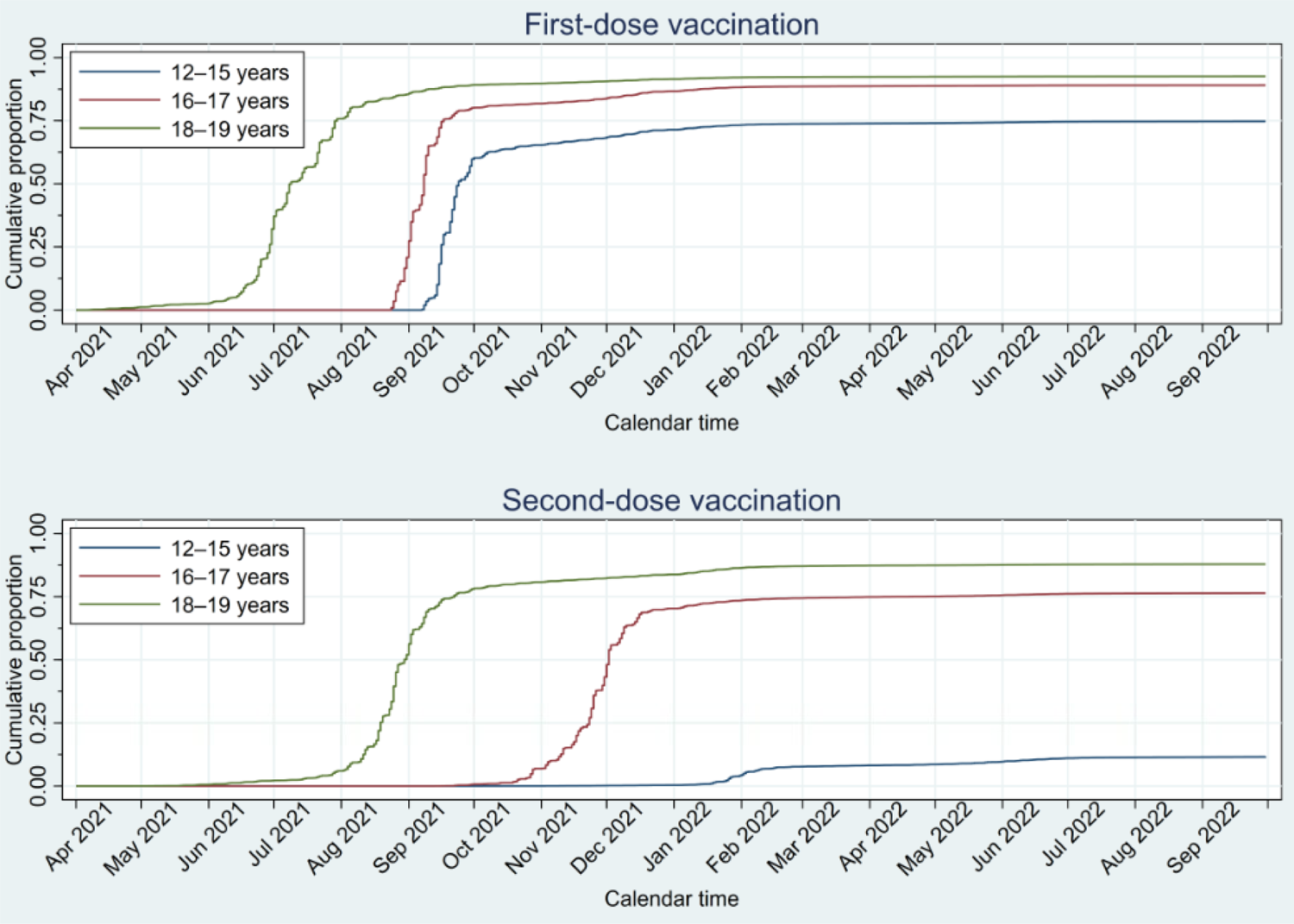
Kaplan-Meier estimate of the cumulative proportion of included study subjects vaccinated against SARS-CoV-2 with an mRNA vaccine, Comirnaty (Pfizer-BioNTech) or Spikevax (Moderna), as a function of calendar time by age group for first-dose vaccination (upper panel) and second-dose vaccination (lower pan

By study end (September 30, 2022), 82.5% had completed first-dose SARS-CoV-2 vaccination (Table 2). Furthermore, 11.5%, 76.4% and 87.2% of subjects aged 12–15, 16–17 and 18–19 years, respectively, had received a second dose.

### Poisson Regression

Results of the main analyses are displayed in Table 3. There were no statistically significant associations between first-dose vaccination and any outcomes inside the risk windows. Second-dose vaccination was associated with anaphylactic reaction (adjusted IRR [aIRR]: 10.05; 95% CI: 1.22– 82.74), which should be interpreted cautiously due to small numbers. We also observed associations with lymphadenopathy, and myocarditis and pericarditis following second-dose vaccination, with aIRRs of 2.33 (95% CI: 1.46–3.72) and 5.27 (95% CI: 1.98–14.05), respectively. Outside the risk windows, we observed statistically significant associations between vaccination and acute appendicitis and facial nerve palsy (Table 3).

**Table 3:**
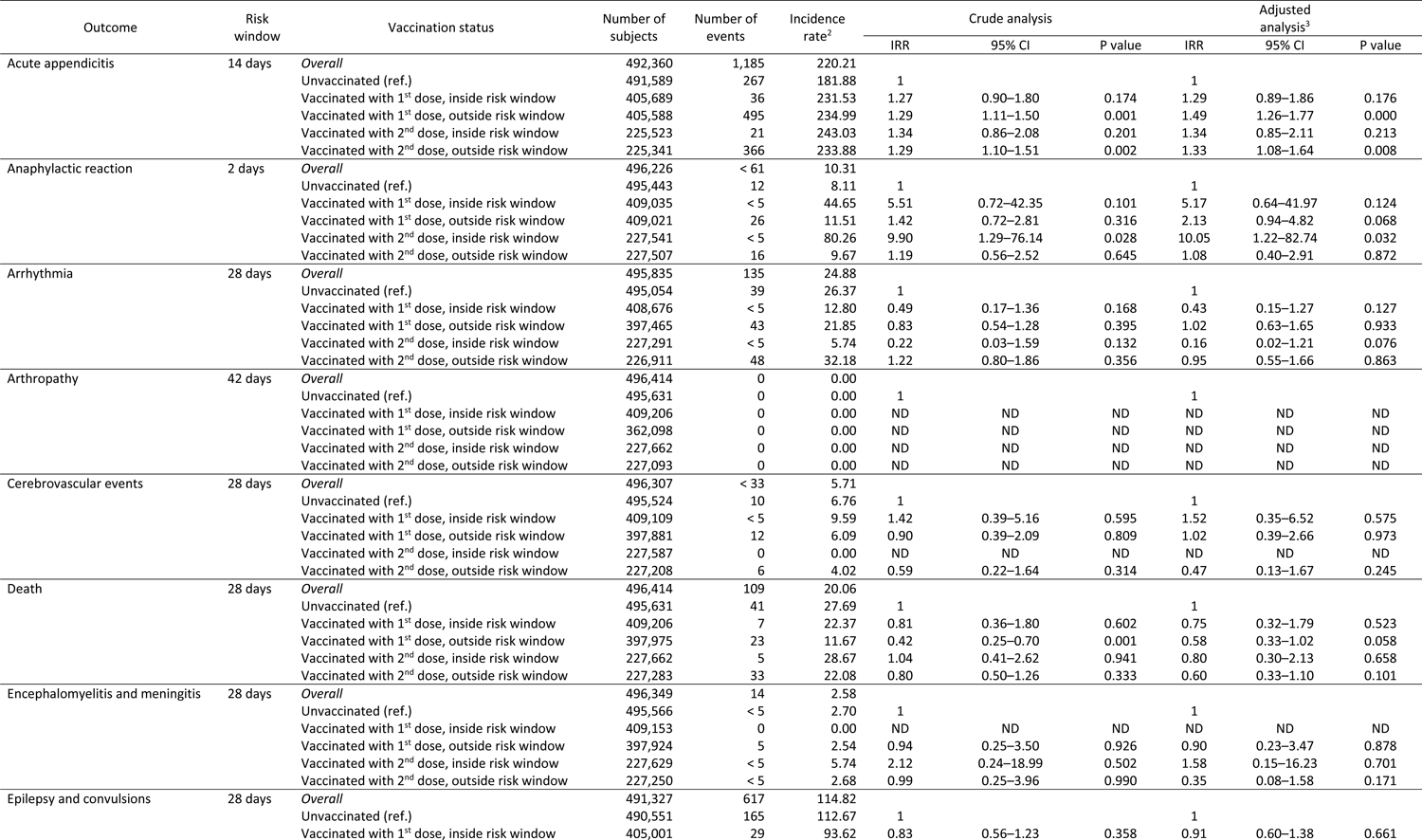
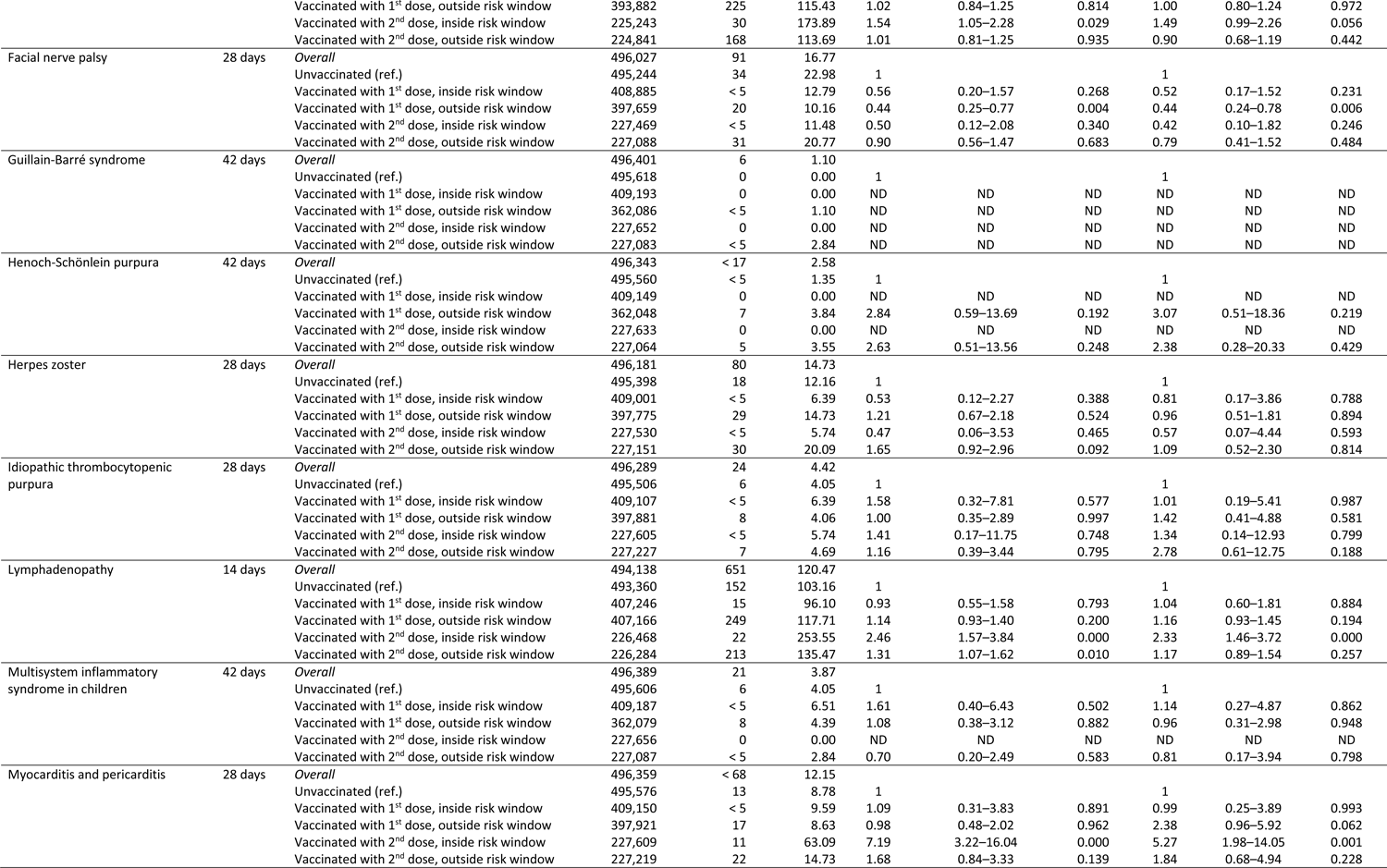
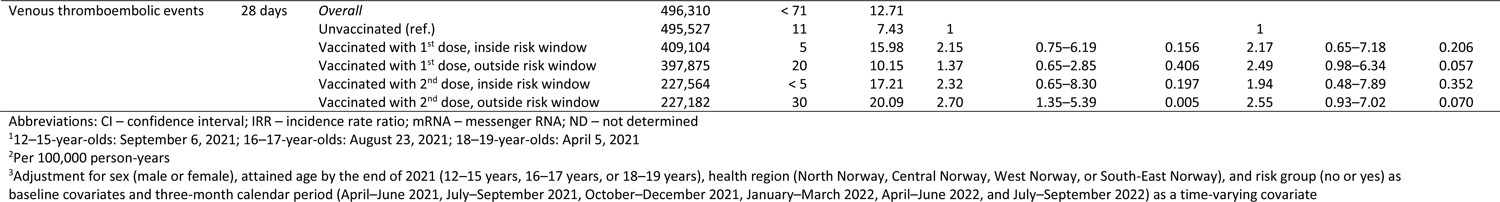
Crude and adjusted incidence rate ratios of 17 different outcomes between vaccinated and unvaccinated subjects, with associated 95% confidence intervals, based on Poisson regression of 496,432 adolescents in Norway aged 12–19 years at the end of 2021 and unvaccinated against SARS-CoV-2 at the beginning of follow-up. Subjects were followed from the beginning of the wave of vaccination^1^ of their age group until the outcome in question, non-mRNA SARS-CoV-2 vaccination, third-dose SARS-CoV-2 vaccination, emigration, death, or end of study on September 30, 2022, whichever occurred first. To ensure data privacy, numbers between 1 and 4 have been suppressed and are denoted by “< 5”. As a result, some of the totals have been suppressed as well to avoid revealing small numbers that have been suppressed.

Supplementary Table 1 displays sensitivity analyses restricted to previously uninfected subjects. Results were similar to the main analyses inside the risk windows, in addition to increased risk of epilepsy and convulsions following second-dose vaccination (aIRR: 1.65; 95% CI: 1.05–2.59). We also observed statistically significant associations between vaccination and acute appendicitis, anaphylactic reaction, death, and myocarditis and pericarditis outside the risk windows (Supplementary Table 1).

### Age-Stratified Analyses

Results of the age-stratified Poisson regression analyses are displayed in Supplementary Tables 2–4. First-dose vaccination in 12–15-year-olds was associated with increased risks of acute appendicitis (aIRR: 1.91; 95% CI: 1.12–3.27) and anaphylactic reaction (aIRR: 16.19; 95% CI: 1.29–202.98) (Supplementary Table 2), although the latter was based on few cases and considered biologically implausible. Furthermore, we observed increased myocarditis and pericarditis risk following second-dose vaccination (aIRR: 37.07; 95% CI: 2.79–492.94) (Supplementary Table 2), but the number of cases was small. In 16–17-year-olds, second-dose vaccination was associated with increased acute appendicitis risk (aIRR: 2.81; 95% CI: 1.30–6.09) (Supplementary Table 3). 18–19-year-olds had increased anaphylactic reaction risk following second-dose vaccination (aIRR: 38.78; 95% CI: 3.46– 434.39) (Supplementary Table 4), but few cases were observed. Second-dose vaccination was also associated with increased risks of epilepsy and convulsions (aIRR: 2.20; 95% CI: 1.05–4.61), lymphadenopathy (aIRR: 2.37; 95% CI: 1.09–5.15), and myocarditis and pericarditis (aIRR: 10.25; 95% CI: 2.36–44.47) (Supplementary Table 4). As in the main analyses, there were also some statistically significant associations outside the risk windows in each age group (Supplementary Tables 2–4).

### Post-hoc Analyses

We observed increased acute appendicitis risk outside the 14-day risk window following both first- and second-dose vaccinations in the main analysis (Table 3). Therefore, we conducted post-hoc analyses using risk windows of 28, 42, and 56 days to explore whether the manifestation of acute appendicitis might be longer than initially presumed.

We observed no statistically significant association between first- or second-dose vaccination and acute appendicitis inside the 28-day risk window. With 42 days there was increased risk following both first (aIRR: 1.39; 95% CI: 1.09–1.78) and second-dose vaccination (aIRR: 1.43; 95% CI: 1.07–1.91). The same was seen when using 56 days, with aIRRs of 1.47 (95% CI: 1.17–1.83) and 1.44 (95% CI: 1.10–1.88), respectively. As data-driven, these results are speculative and must be interpreted cautiously.

### SCCS Analysis

Supplementary Table 5 displays results of the SCCS analysis. We observed no statistically significant increase in risk associated with first-dose vaccination for any outcomes inside the risk windows. Like in the Poisson regression analyses, second-dose vaccination was associated with lymphadenopathy (aIRR: 2.04; 95% CI: 1.24–3.35) and myocarditis and pericarditis (aIRR: 5.88; 95% CI: 2.11–16.40).

Furthermore, we observed a borderline statistically significant association between second-dose vaccination and epilepsy and convulsions (aIRR: 1.59; 95% CI: 0.99–2.55), and herpes zoster outside the second risk window (aIRR: 4.12: 95%CI: 1.10–15.41). There was also a tendency towards increased anaphylactic reaction risk following second-dose vaccination, but this association was based on few cases.

In the sensitivity analysis including a 14-day preexposure risk period, results remained unchanged for most outcomes. Associations of second-dose vaccination with lymphadenopathy and myocarditis and pericarditis were slightly attenuated (aIRRs 1.78 [95% CI: 1.06–2.97] and 4.55 [95% CI: 1.62– 12.68], respectively), whereas the associations with herpes zoster, epilepsy and convulsions, lost statistical significance.

## Discussion

We investigated 17 preselected potential adverse events following SARS-CoV-2 mRNA vaccination in adolescents aged 12–19 years in Norway. The main analyses showed no statistically significant associations between first-dose vaccination and any outcomes, whereas second-dose vaccination was associated with anaphylactic reaction, lymphadenopathy, and myocarditis and pericarditis.

### Research in Context

There are case reports/series for most outcomes investigated,[15–26] possibly due to random co-occurrences. Few adolescent studies are currently published. Lai et al. investigated post-vaccination adverse events in adolescents,[27] but differences in ages, risk windows, and diagnostic coding in addition to national differences in policy, infection, and vaccination rates during the COVID-19 pandemic make comparisons complex.

#### Acute Appendicitis

A nationwide Israeli study reported increased risk,[10] while nationwide Danish and Swedish studies did not.[28, 29] In our main analyses, we found no associations inside the risk windows. In age-stratified analyses, we observed increased risk following first-dose vaccination in 12–15-year-olds and second-dose vaccination in 16–17-year-olds. These findings must be interpreted cautiously, but do not necessarily conflict with Scandinavian results in older individuals (12–24 years).[28, 29]

#### Anaphylactic Reaction

Anaphylactic reaction was rare, with <5 cases after both doses. Contrary to Lai et al.,[27] we observed increased risk following second-dose vaccination. We included only inpatient care, while Lai et al. included both in- and outpatient care.[27]

#### Death

We found no statistically significant associations with all-cause mortality within 28 days. Events were very rare. No Norwegian adolescents were registered with vaccine-associated death (International Statistical Classification of Diseases and Related Health Problems, Tenth Revision [ICD-10] code U12.9) during follow-up.

#### Epilepsy and Convulsions

Adult studies are inconsistent.[30–33] Lai et al. found no statistically significant association.[27] We observed increased risk following second-dose vaccination in uninfected subjects and 18–19-year-olds using Poisson regression. Uninfected subjects could be overrepresented by subjects with increased risk of epilepsy and convulsions, who presumably adhered more strictly to protective measures. The SCCS analysis showed a potential association with second-dose vaccination, which disappeared when including a 14-day preexposure period. These results should be interpreted very cautiously.

#### Facial Nerve Palsy

Data from the Pfizer clinical trial reported four vaccinees with Bell’s palsy vs. none in the placebo group,[4] but other studies report no statistically significant association,[10, 30, 34–37] supporting our results.

#### Herpes Zoster

Some studies report increased post-vaccination herpes zoster risk in adults and adolescent males,[10, 38] whereas others do not.[36] We found no consistent statistically significant association, but few cases were observed.

#### Lymphadenopathy

Lymphadenopathy is a very common post-vaccination event. More lymphadenopathy cases were reported in vaccinees in two multinational clinical trials and a nationwide Israeli adolescent study,[6, 8, 10] supporting our results.

#### Myocarditis and Pericarditis

A previous Nordic study reported increased risk in ages ≥12 years,[39] as have adolescent studies,[10, 27, 40] particularly after the second dose.[41] This was not observed in clinical trials,[5, 7–9] nor in a Malaysian study.[30] Our results support increased risk after vaccination.

#### Arrythmia

Myocarditis may lead to arrhythmia, but we observed no vaccine-arrhythmia association. Other studies reporting increased post-vaccination myocarditis risk similarly report no vaccine-arrhythmia association,[10, 27, 42] while a Malaysian study report increased risk.[30] Post-vaccination myocarditis might be milder,[40, 43] resulting in fewer arrhythmias, or arrhythmias might be less common in adolescents.

#### Cerebrovascular Events and Venous Thromboembolic Events

Cerebrovascular events were grouped in the current study. While individually rare in adolescents, they could be considered different manifestations of a common pathology. Similar reasoning was used when grouping venous thromboembolic events.

Some adult studies report increased risks of venous thromboembolic events and hemorrhagic events following vaccination,[30, 44] whereas others do not.[45–47] A Nordic adult study found no consistent associations with cerebrovascular disease or coagulation disorders,[48] whereas an adolescent study reported no associations with thromboembolic or cerebrovascular events (excepting myocarditis).[27] A study in 16–19-year-olds reported no associations between vaccination and deep vein thrombosis or pulmonary embolism.[38]

We found no statistically significant associations between vaccination and cerebrovascular events or venous thromboembolic events. Estimates of venous thromboembolic events were elevated following first- and second-dose vaccinations, both inside and outside risk windows in the Poisson analysis (Table 3). These events should therefore be further investigated.

#### Other Adverse Events

Although most outcomes studied were not significantly associated with vaccination, some (arrhythmia, arthropathy, cerebrovascular events, encephalomyelitis and meningitis, Guillain-Barré syndrome, Henoch-Schönlein purpura, idiopathic thrombocytopenic purpura, and multisystem inflammatory syndrome in children [MIS-C]) cannot be ruled out due to rarity.

We observed no arthropathy cases. Arthropathy is rare and may develop slowly in adolescents, necessitating longer follow-up. Post-vaccination Henoch-Schönlein purpura has been proposed,[25, 49] but we observed no events inside the risk windows in our study. Lai et al. reported few or no cases of idiopathic thrombocytopenia, Guillain-Barré syndrome, meningoencephalitis, and MIS-C and no vaccine associations.[27] MIS-C has been estimated at 1/1,000,000 vaccinees,[22] which makes our study underpowered. Similarly, other studies generally report few events or null associations.[31, 35–37, 46, 47, 50]

### Strengths and Limitations

The main strength was individual-level data from high-quality nationwide health registers with mandatory reporting. Norwegian health care and SARS-CoV-2 vaccination is nominally free of charge, reducing socioeconomic confounding. By applying different methods, we could check the robustness of our results. We adjusted for potential confounders in the Poisson regression analysis, while time-invariant confounders were implicitly adjusted for in the SCCS analysis.[13, 14] High vaccination coverage means adverse events should be rare or weakly associated to remain undetected.

We excluded subjects vaccinated prior to their age-specific vaccination wave, as off-label use was generally reserved for high-risk groups. Although media attention might have introduced some vigilance or notoriety bias, limiting outcomes to diagnoses from hospital contacts for all outcomes except for herpes zoster likely limited outcomes to the more severe cases. A study period of 18 months means that the study was not limited to early vaccinees, who might constitute a selected population (e.g., underlying diseases or high socioeconomic status). As there was no strong recommendation in Norway to vaccinate healthy adolescents <16 years old, the unvaccinated reference group was less selected and more representative of the population. Adjusting for calendar time lessened the risk of temporal associations influencing results, mitigating effects of lifestyle changes, pandemic restrictions, or other unmeasured factors correlated with time.

Our main limitation was rare outcomes leading to unreliable estimates. There might be confounding by indication and unmeasured confounding, as vaccinated and unvaccinated subjects might differ in health status and health-seeking behavior, e.g., lower threshold for medical consultation among vaccinees. Conversely, there might be a healthy-vaccinee effect, where unwell subjects forego vaccination. Lifestyle-related and societal changes in physical activity, diet, and pandemic restrictions during follow-up might have been present, but adolescents were vaccinated when restrictions were few. The SCCS preexposure risk period for second-dose vaccination might fall inside the first-dose risk window. E.g., if the first and second doses were administered within 28 days and the risk window was 28 days, the second-dose 14-day preexposure period would limit the risk window following first-dose vaccination to 14 days. This was only relevant for 18–19-year-olds, with recommended minimum dose interval of 6–12 weeks.

### Conclusion and Future Research

Knowledge of potential post-vaccination adverse events is crucial to weigh benefits against risks, and for future vaccine recommendations. The number of observed outcomes and statistically significant associations were generally low in this study, with some exceptions which should be further monitored. More adolescent studies are necessary to explore potential age-specific post-vaccination adverse events, especially in relation to new mRNA vaccines or boosters.

## Data Availability

Due to the nature of the Emergency register, the authors do not have permission to share individual-level data. Data are only accessible to authorized researchers pending ethical approval and successful application to www.helsedata.no.

## Author contributions

Vilde Bergstad Larsen: Methodology, Software, Formal Analysis, Writing-Original Draft, Review & Editing, Visualization.

Nina Gunnes: Conceptualization, Methodology, Software, Formal Analysis, Data Curation, Writing-Original Draft, Review & Editing, Visualization.

Jon Michael Gran: Methodology, Writing – Review & Editing.

Jesper Dahl: Conceptualization, Data Curation, Writing – Review & Editing.

Håkon Bøås: Conceptualization, Data Curation, Writing – Review & Editing.

Sara Viksmoen Watle: Conceptualization, Writing – Review & Editing.

Jacob Dag Berild: Conceptualization, Writing – Review & Editing.

Margrethe Greve-Isdahl: Conceptualization, Writing – Review & Editing.

Ketil Størdal: Conceptualization, Writing – Review & Editing.

Hanne Løvdal Gulseth: Conceptualization, Writing – Review & Editing, Project administration, Funding Acquisition.

Øystein Karlstad: Conceptualization, Methodology, Writing – Review & Editing.

Paz Lopez-Doriga Ruiz: Conceptualization, Data Curation, Writing – Review & Editing.

German Tapia: Conceptualization, Validation, Data Curation, Writing - Original Draft, Review & Editing, Supervision, Project administration.

Conflict of interest: ØK and PLDR reports participation in research projects funded by Novo Nordisk, LEO Pharma and Bristol-Myers Squibb, all regulator-mandated phase IV studies, all with funds paid to their institution (no personal fees) and with no relation to the work reported in this paper. HLG reports previous participation in research projects and clinical trials funded by Novo Nordisk, GSK, AstraZeneca, and Boehringer-Ingelheim, all related to diabetes and paid to her previous institution Oslo University Hospital (no personal fees). She has received a lecture fee from Sanofi-Aventis (2018). All other authors report no conflict of interest.

Ethics: The emergency preparedness register Beredt C19 was established by the Norwegian Institute of Public Health according to the Health Preparedness Act §2-4, and the study was approved by the Regional Committee for Medical and Health Research Ethics South-East Norway (REK Sør-Øst A, ref 122745).

Data availability: Due to the nature of the Emergency register, the authors do not have permission to share individual-level data. Data are only accessible to authorized researchers pending ethical approval and successful application to www.helsedata.no.

Funding: None.

