## Supplemental material for "Adverse Events Following SARS-CoV-2 mRNA Vaccination in Adolescents: A Norwegian Nationwide Register-Based Study"

Vilde Bergstad Larsen[1], Nina Gunnes[2,3], Jon Michael Gran[4], Jesper Dahl[5], Håkon Bøås[5], Sara Viksmoen Watle[5], Jacob Dag Berild[5], Margrethe Greve-Isdahl[5], Ketil Størdal[6], Hanne Løvdal Gulseth[2], Øystein Karlstad[2], Paz Lopez-Doriga Ruiz[2,7], German Tapia[2]

[1]Division for Health Services, Norwegian Institute of Public Health, Oslo, Norway
[2]Division of Mental and Physical Health, Norwegian Institute of Public Health, Oslo, Norway
[3]Norwegian Research Centre for Women's Health, Oslo University Hospital, Oslo, Norway
[4]Oslo Centre for Biostatistics and Epidemiology, Department of Biostatistics, University of Oslo, Oslo, Norway
[5]Department of Infection Control and Vaccines, Norwegian Institute of Public Health, Oslo, Norway
[6]Department of Pediatric Research, Institute of Clinical Medicine, University of Oslo, and Oslo University Hospital, Oslo, Norway
[7]Institute of Community Health and Global Medicine, University of Oslo, Oslo, Norway

### Supplementary Materials

This section includes additional information about the current study.

#### Data Sources in Beredt C19

Beredt C19 was established in April 2020 by the Norwegian Institute of Public Health to monitor infections, vaccinations, and use of health services in Norway during the COVID-19 pandemic (https://www.fhi.no/en/id/infectious-diseases/coronavirus/emergency-preparedness-register-for-covid-19/). The register consists of historical and real-time data from various nationwide electronic health registers and administrative databases to which all reporting is mandatory, thereby covering the entire population in Norway. Data sources in Beredt C19 relevant for the current study are described below.

##### National Population Register

The National Population Register (https://www.skatteetaten.no/en/person/national-registry/) provided information on date and place of birth, sex, date of immigration, county of residence, date of emigration, date of death, and residential status (resident, emigrated, or deceased). The register is updated weekly in Beredt C19.

##### Norwegian Surveillance System for Communicable Diseases

Information on laboratory-confirmed SARS-CoV-2 infection dates was provided by the Norwegian Surveillance System for Communicable Diseases (MSIS), which is run by the Norwegian Institute of Public Health. All laboratories are legally required to report date of testing and test results to the MSIS. The register is updated daily in Beredt C19.

##### Norwegian Immunisation Registry

The Norwegian Immunisation Registry (SYSVAK) provided information on SARS-CoV-2 vaccination dates and vaccine types. SYSVAK is run by the Norwegian Institute of Health and updated daily in Beredt C19.

##### Norwegian Registry of Primary Health Care

The Norwegian Registry of Primary Health Care is based on government reimbursement claims from the primary health services in Norway, including publicly funded general practitioners and primary care emergency clinics. Diagnoses are coded using the International Classification of Primary Care, Second Edition (ICPC-2). The register is updated daily in Beredt C19. In the current study, it provided information on date of diagnosis of herpes zoster (ICPC-2 code S70).

##### Norwegian Patient Registry

The Norwegian Patient Registry (NPR) collects data from the specialist health services in Norway, which includes all government-owned hospitals and outpatient clinics. Diagnoses are coded using the International Statistical Classification of Diseases and Related Health Problems, Tenth Revision (ICD-10). Reporting to the NPR forms the basis for government reimbursements to the specialist health services in Norway. Together with the Norwegian Registry of Primary Health Care, the NPR covers all government-funded health care in Norway. The register is updated daily in Beredt C19. It provided information on dates of all hospital admissions/discharges and outpatient visits and the corresponding diagnoses.

##### Norwegian Cause of Death Registry

Information on county of residence for deceased subjects was retrieved from the Norwegian Cause of Death Registry, as home address is deleted from the National Population Register upon death. We also used the Norwegian Cause of Death Registry to ascertain whether any subjects were registered with vaccine-associated death (ICD-10 code U12.9). The register is run by the Norwegian Institute of Public Health and updated weekly in Beredt C19.

*Supplementary Table 1: Crude and adjusted incidence rate ratios of 17 different outcomes between vaccinated and unvaccinated subjects, with associated 95% confidence intervals, based on Poisson regression of 477,097 adolescents in Norway aged 12–19 years at the end of 2021 and unvaccinated against SARS-CoV-2 and previously uninfected with SARS-CoV-2 at the beginning of follow-up. Subjects were followed from the beginning of the wave of vaccination[1] of their age group until the outcome in question, non-mRNA SARS-CoV-2 vaccination, third-dose SARS-CoV-2 vaccination, SARS-CoV-2 infection, emigration, death, or end of study on September 30, 2022, whichever occurred first. To ensure data privacy, numbers between 1 and 4 have been suppressed and are denoted by "< 5". As a result, some of the totals have been suppressed as well to avoid revealing small numbers that have been suppressed.*

| Outcome | Risk window | Vaccination status | Number of subjects | Number of events | Incidence rate[2] | Crude analysis | | | Adjusted analysis[3] | | |
|---|---|---|---|---|---|---|---|---|---|---|---|
| | | | | | | IRR | 95% CI | P value | IRR | 95% CI | P value |
| Acute appendicitis | 14 days | *Overall* | 472,625 | 802 | 217.77 | | | | | | |
| | | Unvaccinated (ref.) | 471,868 | 184 | 184.88 | 1 | | | 1 | | |
| | | Vaccinated with 1st dose, inside risk window | 388,948 | 34 | 229.38 | 1.24 | 0.86–1.79 | 0.248 | 1.33 | 0.90–1.96 | 0.147 |
| | | Vaccinated with 1st dose, outside risk window | 384,298 | 300 | 229.31 | 1.24 | 1.03–1.49 | 0.021 | 1.56 | 1.25–1.94 | 0.000 |
| | | Vaccinated with 2nd dose, inside risk window | 213,406 | 20 | 246.64 | 1.33 | 0.84–2.12 | 0.221 | 1.42 | 0.87–2.30 | 0.159 |
| | | Vaccinated with 2nd dose, outside risk window | 210,127 | 264 | 229.58 | 1.24 | 1.03–1.50 | 0.024 | 1.45 | 1.10–1.90 | 0.008 |
| Anaphylactic reaction | 2 days | *Overall* | 476,333 | 41 | 11.04 | | | | | | |
| | | Unvaccinated (ref.) | 475,564 | 9 | 8.97 | 1 | | | 1 | | |
| | | Vaccinated with 1st dose, inside risk window | 392,153 | < 5 | 46.58 | 5.19 | 0.66–40.98 | 0.118 | 6.84 | 0.81–57.81 | 0.077 |
| | | Vaccinated with 1st dose, outside risk window | 391,666 | 19 | 13.13 | 1.46 | 0.66–3.23 | 0.347 | 2.75 | 1.00–7.56 | 0.049 |
| | | Vaccinated with 2nd dose, inside risk window | 215,309 | < 5 | 84.86 | 9.46 | 1.20–74.65 | 0.033 | 11.02 | 1.26–96.68 | 0.030 |
| | | Vaccinated with 2nd dose, outside risk window | 214,849 | 11 | 8.94 | 1.00 | 0.41–2.40 | 0.993 | 0.88 | 0.26–2.95 | 0.830 |
| Arrhythmia | 28 days | *Overall* | 475,961 | 90 | 24.25 | | | | | | |
| | | Unvaccinated (ref.) | 475,194 | 26 | 25.94 | 1 | | | 1 | | |
| | | Vaccinated with 1st dose, inside risk window | 391,814 | < 5 | 10.11 | 0.39 | 0.12–1.29 | 0.122 | 0.38 | 0.11–1.31 | 0.126 |
| | | Vaccinated with 1st dose, outside risk window | 374,429 | 24 | 20.49 | 0.79 | 0.45–1.38 | 0.405 | 1.12 | 0.58–2.16 | 0.726 |
| | | Vaccinated with 2nd dose, inside risk window | 215,069 | < 5 | 6.15 | 0.24 | 0.03–1.75 | 0.158 | 0.20 | 0.03–1.56 | 0.126 |
| | | Vaccinated with 2nd dose, outside risk window | 210,483 | 36 | 33.37 | 1.29 | 0.78–2.13 | 0.328 | 1.34 | 0.64–2.82 | 0.438 |
| Arthropathy | 42 days | *Overall* | 476,515 | 0 | 0.00 | | | | | | |
| | | Unvaccinated (ref.) | 475,746 | 0 | 0.00 | 1 | | | 1 | | |
| | | Vaccinated with 1st dose, inside risk window | 392,319 | 0 | 0.00 | ND | ND | ND | ND | ND | ND |
| | | Vaccinated with 1st dose, outside risk window | 336,587 | 0 | 0.00 | ND | ND | ND | ND | ND | ND |
| | | Vaccinated with 2nd dose, inside risk window | 215,425 | 0 | 0.00 | ND | ND | ND | ND | ND | ND |
| | | Vaccinated with 2nd dose, outside risk window | 208,863 | 0 | 0.00 | ND | ND | ND | ND | ND | ND |
| Cerebrovascular events | 28 days | *Overall* | 476,409 | 22 | 5.92 | | | | | | |
| | | Unvaccinated (ref.) | 475,640 | 8 | 7.97 | 1 | | | 1 | | |
| | | Vaccinated with 1st dose, inside risk window | 392,224 | < 5 | 10.10 | 1.27 | 0.34–4.77 | 0.727 | 1.26 | 0.28–5.58 | 0.764 |
| | | Vaccinated with 1st dose, outside risk window | 374,812 | 7 | 5.97 | 0.75 | 0.27–2.07 | 0.577 | 0.88 | 0.26–2.98 | 0.838 |
| | | Vaccinated with 2nd dose, inside risk window | 215,353 | 0 | 0.00 | ND | ND | ND | ND | ND | ND |
| | | Vaccinated with 2nd dose, outside risk window | 210,758 | < 5 | 3.70 | 0.46 | 0.14–1.54 | 0.210 | 0.27 | 0.06–1.28 | 0.100 |
| Death | 28 days | *Overall* | 476,515 | 90 | 24.22 | | | | | | |
| | | Unvaccinated (ref.) | 475,746 | 36 | 35.87 | 1 | | | 1 | | |
| | | Vaccinated with 1st dose, inside risk window | 392,319 | 7 | 23.56 | 0.66 | 0.29–1.48 | 0.309 | 0.71 | 0.29–1.69 | 0.433 |
| | | Vaccinated with 1st dose, outside risk window | 374,903 | 18 | 15.35 | 0.43 | 0.24–0.75 | 0.003 | 0.59 | 0.31–1.13 | 0.110 |
| | | Vaccinated with 2nd dose, inside risk window | 215,425 | 5 | 30.68 | 0.86 | 0.34–2.18 | 0.743 | 0.69 | 0.26–1.87 | 0.471 |
| | | Vaccinated with 2nd dose, outside risk window | 210,830 | 24 | 22.21 | 0.62 | 0.37–1.04 | 0.069 | 0.40 | 0.20–0.78 | 0.007 |
| Encephalomyelitis and meningitis | 28 days | *Overall* | 476,453 | 7 | 1.88 | | | | | | |
| | | Unvaccinated (ref.) | 475,684 | < 5 | 1.00 | 1 | | | 1 | | |
| | | Vaccinated with 1st dose, inside risk window | 392,267 | 0 | 0.00 | ND | ND | ND | ND | ND | ND |
| | | Vaccinated with 1st dose, outside risk window | 374,854 | < 5 | 1.71 | 1.71 | 0.16–18.88 | 0.661 | 1.18 | 0.10–13.79 | 0.895 |
| | | Vaccinated with 2nd dose, inside risk window | 215,394 | < 5 | 6.14 | 6.16 | 0.39–98.45 | 0.199 | 5.98 | 0.28–126.35 | 0.251 |
| | | Vaccinated with 2nd dose, outside risk window | 210,799 | < 5 | 2.78 | 2.79 | 0.29–26.78 | 0.375 | 1.69 | 0.11–26.81 | 0.709 |
| Epilepsy and convulsions | 28 days | *Overall* | 471,600 | 454 | 123.56 | | | | | | |

| | | | | | | | | | | | |
|---|---|---|---|---|---|---|---|---|---|---|---|
| | | Unvaccinated (ref.) | 470,838 | 107 | 107.85 | 1 | | | 1 | | |
| | | Vaccinated with 1st dose, inside risk window | 388,243 | 29 | 98.63 | 0.91 | 0.61–1.38 | 0.670 | 1.07 | 0.69–1.67 | 0.749 |
| | | Vaccinated with 1st dose, outside risk window | 370,996 | 157 | 135.46 | 1.26 | 0.98–1.61 | 0.069 | 1.21 | 0.91–1.61 | 0.187 |
| | | Vaccinated with 2nd dose, inside risk window | 213,099 | 28 | 173.71 | 1.61 | 1.06–2.44 | 0.025 | 1.65 | 1.05–2.59 | 0.028 |
| | | Vaccinated with 2nd dose, outside risk window | 208,521 | 133 | 124.53 | 1.15 | 0.90–1.49 | 0.268 | 1.01 | 0.71–1.44 | 0.944 |
| Facial nerve palsy | 28 days | *Overall* | 476,148 | 57 | 15.35 | | | | | | |
| | | Unvaccinated (ref.) | 475,379 | 19 | 18.95 | 1 | | | 1 | | |
| | | Vaccinated with 1st dose, inside risk window | 392,016 | < 5 | 13.47 | 0.71 | 0.24–2.09 | 0.535 | 0.65 | 0.21–2.02 | 0.454 |
| | | Vaccinated with 1st dose, outside risk window | 374,613 | 10 | 8.54 | 0.45 | 0.21–0.97 | 0.041 | 0.47 | 0.20–1.11 | 0.085 |
| | | Vaccinated with 2nd dose, inside risk window | 215,248 | < 5 | 12.28 | 0.65 | 0.15–2.78 | 0.560 | 0.49 | 0.11–2.26 | 0.359 |
| | | Vaccinated with 2nd dose, outside risk window | 210,657 | 22 | 20.38 | 1.08 | 0.58–1.99 | 0.816 | 1.02 | 0.41–2.53 | 0.964 |
| Guillain-Barré syndrome | 42 days | *Overall* | 476,503 | < 5 | 0.81 | | | | | | |
| | | Unvaccinated (ref.) | 475,734 | 0 | 0.00 | 1 | | | 1 | | |
| | | Vaccinated with 1st dose, inside risk window | 392,307 | 0 | 0.00 | ND | ND | ND | ND | ND | ND |
| | | Vaccinated with 1st dose, outside risk window | 336,577 | 0 | 0.00 | ND | ND | ND | ND | ND | ND |
| | | Vaccinated with 2nd dose, inside risk window | 215,416 | 0 | 0.00 | ND | ND | ND | ND | ND | ND |
| | | Vaccinated with 2nd dose, outside risk window | 208,855 | < 5 | 3.00 | ND | ND | ND | ND | ND | ND |
| Henoch-Schönlein purpura | 42 days | *Overall* | 476,444 | 7 | 1.88 | | | | | | |
| | | Unvaccinated (ref.) | 475,675 | < 5 | 1.00 | 1 | | | 1 | | |
| | | Vaccinated with 1st dose, inside risk window | 392,263 | 0 | 0.00 | ND | ND | ND | ND | ND | ND |
| | | Vaccinated with 1st dose, outside risk window | 336,538 | 5 | 4.84 | 4.85 | 0.57–41.52 | 0.149 | ND | ND | ND |
| | | Vaccinated with 2nd dose, inside risk window | 215,397 | 0 | 0.00 | ND | ND | ND | ND | ND | ND |
| | | Vaccinated with 2nd dose, outside risk window | 208,835 | < 5 | 1.00 | 1.00 | 0.06–16.04 | 0.998 | ND | ND | ND |
| Herpes zoster | 28 days | *Overall* | 476,297 | 47 | 12.65 | | | | | | |
| | | Unvaccinated (ref.) | 475,528 | 9 | 8.97 | 1 | | | 1 | | |
| | | Vaccinated with 1st dose, inside risk window | 392,127 | < 5 | 6.73 | 0.75 | 0.16–3.47 | 0.714 | 1.00 | 0.20–5.09 | 0.997 |
| | | Vaccinated with 1st dose, outside risk window | 374,721 | 15 | 12.80 | 1.43 | 0.62–3.26 | 0.399 | 1.30 | 0.49–3.42 | 0.598 |
| | | Vaccinated with 2nd dose, inside risk window | 215,299 | < 5 | 6.14 | 0.68 | 0.09–5.40 | 0.719 | 0.82 | 0.10–6.85 | 0.853 |
| | | Vaccinated with 2nd dose, outside risk window | 210,710 | 20 | 18.52 | 2.06 | 0.94–4.53 | 0.071 | 1.86 | 0.62–5.56 | 0.265 |
| Idiopathic thrombocytopenic purpura | 28 days | *Overall* | 476,398 | 18 | 4.84 | | | | | | |
| | | Unvaccinated (ref.) | 475,629 | 5 | 4.98 | 1 | | | 1 | | |
| | | Vaccinated with 1st dose, inside risk window | 392,227 | < 5 | 6.73 | 1.35 | 0.26–6.96 | 0.719 | 0.98 | 0.18–5.44 | 0.983 |
| | | Vaccinated with 1st dose, outside risk window | 374,817 | 5 | 4.27 | 0.86 | 0.25–2.96 | 0.806 | 1.28 | 0.29–5.71 | 0.748 |
| | | Vaccinated with 2nd dose, inside risk window | 215,372 | < 5 | 6.14 | 1.23 | 0.14–10.54 | 0.849 | 1.16 | 0.11–11.87 | 0.901 |
| | | Vaccinated with 2nd dose, outside risk window | 210,779 | 5 | 4.63 | 0.93 | 0.27–3.21 | 0.907 | 2.18 | 0.37–12.99 | 0.391 |
| Lymphadenopathy | 14 days | *Overall* | 474,345 | 438 | 118.44 | | | | | | |
| | | Unvaccinated (ref.) | 473,581 | 97 | 97.10 | 1 | | | 1 | | |
| | | Vaccinated with 1st dose, inside risk window | 390,457 | 15 | 100.80 | 1.04 | 0.60–1.79 | 0.893 | 1.23 | 0.70–2.18 | 0.474 |
| | | Vaccinated with 1st dose, outside risk window | 385,807 | 157 | 119.52 | 1.23 | 0.96–1.59 | 0.108 | 1.29 | 0.96–1.73 | 0.094 |
| | | Vaccinated with 2nd dose, inside risk window | 214,303 | 20 | 245.61 | 2.53 | 1.56–4.09 | 0.000 | 2.73 | 1.63–4.58 | 0.000 |
| | | Vaccinated with 2nd dose, outside risk window | 211,014 | 149 | 128.98 | 1.33 | 1.03–1.72 | 0.030 | 1.40 | 0.98–2.01 | 0.068 |
| Multisystem inflammatory syndrome in children | 42 days | *Overall* | 476,500 | 6 | 1.61 | | | | | | |
| | | Unvaccinated (ref.) | 475,731 | < 5 | 1.00 | 1 | | | 1 | | |
| | | Vaccinated with 1st dose, inside risk window | 392,305 | < 5 | 2.30 | 2.30 | 0.14–36.83 | 0.555 | 1.61 | 0.09–28.39 | 0.744 |
| | | Vaccinated with 1st dose, outside risk window | 336,576 | < 5 | 1.93 | 1.94 | 0.18–21.41 | 0.588 | 3.26 | 0.11–93.03 | 0.490 |
| | | Vaccinated with 2nd dose, inside risk window | 215,420 | 0 | 0.00 | ND | ND | ND | ND | ND | ND |
| | | Vaccinated with 2nd dose, outside risk window | 208,858 | < 5 | 2.00 | 2.01 | 0.18–22.13 | 0.570 | 3.73 | 0.08–169.70 | 0.499 |
| Myocarditis and pericarditis | 28 days | *Overall* | 476,463 | < 53 | 13.73 | | | | | | |
| | | Unvaccinated (ref.) | 475,694 | 9 | 8.97 | 1 | | | 1 | | |
| | | Vaccinated with 1st dose, inside risk window | 392,268 | < 5 | 10.10 | 1.13 | 0.30–4.16 | 0.859 | 1.57 | 0.35–7.04 | 0.557 |
| | | Vaccinated with 1st dose, outside risk window | 374,855 | 13 | 11.09 | 1.24 | 0.53–2.89 | 0.625 | 4.84 | 1.42–16.47 | 0.012 |

| | | | | | | | | | | | |
|---|---|---|---|---|---|---|---|---|---|---|---|
| | | Vaccinated with 2nd dose, inside risk window | 215,375 | 11 | 67.52 | 7.53 | 3.12–18.17 | 0.000 | 9.19 | 2.69–31.45 | 0.000 |
| | | Vaccinated with 2nd dose, outside risk window | 210,769 | 15 | 13.89 | 1.55 | 0.68–3.54 | 0.300 | 3.16 | 0.79–12.64 | 0.104 |
| Venous thromboembolic events | 28 days | *Overall* | 476,415 | < 54 | 14.00 | | | | | | |
| | | Unvaccinated (ref.) | 475,646 | 9 | 8.97 | 1 | | | 1 | | |
| | | Vaccinated with 1st dose, inside risk window | 392,221 | 5 | 16.83 | 1.88 | 0.63–5.60 | 0.259 | 2.40 | 0.67–8.62 | 0.180 |
| | | Vaccinated with 1st dose, outside risk window | 374,811 | 13 | 11.09 | 1.24 | 0.53–2.89 | 0.625 | 2.67 | 0.85–8.41 | 0.094 |
| | | Vaccinated with 2nd dose, inside risk window | 215,335 | < 5 | 18.42 | 2.05 | 0.56–7.58 | 0.281 | 2.17 | 0.48–9.73 | 0.313 |
| | | Vaccinated with 2nd dose, outside risk window | 210,737 | 22 | 20.37 | 2.27 | 1.05–4.93 | 0.038 | 2.53 | 0.72–8.85 | 0.147 |

Abbreviations: CI – confidence interval; IRR – incidence rate ratio; mRNA – messenger RNA; ND – not determined

[1]12–15-year-olds: September 6, 2021; 16–17-year-olds: August 23, 2021; 18–19-year-olds: April 5, 2021

[2]Per 100,000 person-years

[3]Adjustment for sex (male or female), attained age by the end of 2021 (12–15 years, 16–17 years, or 18–19 years), health region (North Norway, Central Norway, West Norway, or South-East Norway), and risk group (no or yes) as baseline covariates and three-month calendar period (April–June 2021, July–September 2021, October–December 2021, January–March 2022, April–June 2022, and July–September 2022) as a time-varying covariate

*Supplementary Table 2: Crude and adjusted incidence rate ratios of 17 different outcomes between vaccinated and unvaccinated subjects, with associated 95% confidence intervals, based on Poisson regression of 253,669 adolescents in Norway aged 12–15 years at the end of 2021 and unvaccinated against SARS-CoV-2 at the beginning of follow-up. Subjects were followed from the beginning of the wave of vaccination[1] of their age group until the outcome in question, non-mRNA SARS-CoV-2 vaccination, third-dose SARS-CoV-2 vaccination, emigration, death, or end of study on September 30, 2022, whichever occurred first. To ensure data privacy, numbers between 1 and 4 have been suppressed and are denoted by "< 5". As a result, some of the totals have been suppressed as well to avoid revealing small numbers that have been suppressed.*

| Outcome | Risk window | Vaccination status | Number of subjects | Number of events | Incidence rate[2] | Crude analysis | | | Adjusted analysis[3] | | |
|---|---|---|---|---|---|---|---|---|---|---|---|
| | | | | | | IRR | 95% CI | P value | IRR | 95% CI | P value |
| Acute appendicitis | 14 days | *Overall* | 251,681 | < 539 | 200.50 | | | | | | |
| | | Unvaccinated (ref.) | 250,982 | 123 | 151.17 | 1 | | | 1 | | |
| | | Vaccinated with 1st dose, inside risk window | 188,097 | 18 | 249.69 | 1.65 | 1.01–2.71 | 0.047 | 1.91 | 1.12–3.27 | 0.017 |
| | | Vaccinated with 1st dose, outside risk window | 188,054 | 367 | 225.83 | 1.49 | 1.22–1.83 | 0.000 | 1.40 | 1.14–1.74 | 0.002 |
| | | Vaccinated with 2nd dose, inside risk window | 28,920 | < 5 | 180.78 | 1.20 | 0.30–4.84 | 0.802 | 1.13 | 0.28–4.61 | 0.860 |
| | | Vaccinated with 2nd dose, outside risk window | 28,813 | 26 | 171.73 | 1.14 | 0.74–1.73 | 0.555 | 1.06 | 0.68–1.63 | 0.806 |
| Anaphylactic reaction | 2 days | *Overall* | 253,579 | 21 | 7.79 | | | | | | |
| | | Unvaccinated (ref.) | 252,869 | 5 | 6.10 | 1 | | | 1 | | |
| | | Vaccinated with 1st dose, inside risk window | 189,544 | < 5 | 96.35 | 15.81 | 1.85–135.31 | 0.012 | 16.19 | 1.29–202.98 | 0.031 |
| | | Vaccinated with 1st dose, outside risk window | 189,535 | 13 | 7.64 | 1.25 | 0.45–3.52 | 0.668 | 1.16 | 0.40–3.38 | 0.779 |
| | | Vaccinated with 2nd dose, inside risk window | 29,154 | 0 | 0.00 | ND | ND | ND | ND | ND | ND |
| | | Vaccinated with 2nd dose, outside risk window | 29,124 | < 5 | 12.32 | 2.02 | 0.39–10.42 | 0.401 | 1.35 | 0.25–7.30 | 0.731 |
| Arrhythmia | 28 days | *Overall* | 253,474 | 49 | 18.18 | | | | | | |
| | | Unvaccinated (ref.) | 252,765 | 14 | 17.08 | 1 | | | 1 | | |
| | | Vaccinated with 1st dose, inside risk window | 189,459 | < 5 | 6.89 | 0.40 | 0.05–3.07 | 0.380 | 0.31 | 0.04–2.40 | 0.261 |
| | | Vaccinated with 1st dose, outside risk window | 189,193 | 29 | 18.52 | 1.08 | 0.57–2.05 | 0.803 | 1.26 | 0.62–2.55 | 0.528 |
| | | Vaccinated with 2nd dose, inside risk window | 29,146 | < 5 | 44.94 | 2.63 | 0.35–20.01 | 0.350 | 3.75 | 0.47–30.02 | 0.213 |
| | | Vaccinated with 2nd dose, outside risk window | 28,909 | < 5 | 28.25 | 1.65 | 0.54–5.03 | 0.374 | 1.89 | 0.57–6.22 | 0.294 |
| Arthropathy | 42 days | *Overall* | 253,666 | 0 | 0.00 | | | | | | |
| | | Unvaccinated (ref.) | 252,956 | 0 | 0.00 | 1 | | | 1 | | |
| | | Vaccinated with 1st dose, inside risk window | 189,617 | 0 | 0.00 | ND | ND | ND | ND | ND | ND |
| | | Vaccinated with 1st dose, outside risk window | 189,147 | 0 | 0.00 | ND | ND | ND | ND | ND | ND |
| | | Vaccinated with 2nd dose, inside risk window | 29,176 | 0 | 0.00 | ND | ND | ND | ND | ND | ND |
| | | Vaccinated with 2nd dose, outside risk window | 28,843 | 0 | 0.00 | ND | ND | ND | ND | ND | ND |
| Cerebrovascular events | 28 days | *Overall* | 253,630 | < 15 | 5.19 | | | | | | |
| | | Unvaccinated (ref.) | 252,920 | < 5 | 4.88 | 1 | | | 1 | | |
| | | Vaccinated with 1st dose, inside risk window | 189,585 | 0 | 0.00 | ND | ND | ND | ND | ND | ND |
| | | Vaccinated with 1st dose, outside risk window | 189,320 | 10 | 6.38 | 1.31 | 0.41–4.17 | 0.649 | 1.28 | 0.35–4.70 | 0.706 |
| | | Vaccinated with 2nd dose, inside risk window | 29,168 | 0 | 0.00 | ND | ND | ND | ND | ND | ND |
| | | Vaccinated with 2nd dose, outside risk window | 28,932 | 0 | 0.00 | ND | ND | ND | ND | ND | ND |
| Death | 28 days | *Overall* | 253,666 | 32 | 11.86 | | | | | | |
| | | Unvaccinated (ref.) | 252,956 | 18 | 21.94 | 1 | | | 1 | | |
| | | Vaccinated with 1st dose, inside risk window | 189,617 | < 5 | 6.88 | 0.31 | 0.04–2.35 | 0.259 | 0.43 | 0.05–3.46 | 0.426 |
| | | Vaccinated with 1st dose, outside risk window | 189,352 | 12 | 7.66 | 0.35 | 0.17–0.72 | 0.005 | 0.31 | 0.15–0.66 | 0.002 |
| | | Vaccinated with 2nd dose, inside risk window | 29,176 | 0 | 0.00 | ND | ND | ND | ND | ND | ND |
| | | Vaccinated with 2nd dose, outside risk window | 28,940 | < 5 | 7.05 | 0.32 | 0.04–2.41 | 0.269 | 0.22 | 0.03–1.66 | 0.140 |
| Encephalomyelitis and meningitis | 28 days | *Overall* | 253,632 | 5 | 1.85 | | | | | | |
| | | Unvaccinated (ref.) | 252,922 | < 5 | 2.44 | 1 | | | 1 | | |
| | | Vaccinated with 1st dose, inside risk window | 189,592 | 0 | 0.00 | ND | ND | ND | ND | ND | ND |
| | | Vaccinated with 1st dose, outside risk window | 189,327 | < 5 | 1.91 | 0.78 | 0.13–4.70 | 0.791 | 0.71 | 0.12–4.27 | 0.708 |
| | | Vaccinated with 2nd dose, inside risk window | 29,169 | 0 | 0.00 | ND | ND | ND | ND | ND | ND |
| | | Vaccinated with 2nd dose, outside risk window | 28,933 | 0 | 0.00 | ND | ND | ND | ND | ND | ND |
| Epilepsy and convulsions | 28 days | *Overall* | 251,005 | < 301 | 111.34 | | | | | | |

| | | | | | | | | | | | |
|---|---|---|---|---|---|---|---|---|---|---|---|
| | | Unvaccinated (ref.) | 250,300 | 92 | 113.45 | 1 | | | 1 | | |
| | | Vaccinated with 1st dose, inside risk window | 187,667 | 18 | 125.16 | 1.10 | 0.67–1.83 | 0.703 | 1.28 | 0.75–2.18 | 0.368 |
| | | Vaccinated with 1st dose, outside risk window | 187,388 | 170 | 109.62 | 0.97 | 0.75–1.25 | 0.790 | 0.91 | 0.70–1.18 | 0.467 |
| | | Vaccinated with 2nd dose, inside risk window | 28,818 | < 5 | 45.45 | 0.40 | 0.06–2.87 | 0.363 | 0.33 | 0.05–2.37 | 0.269 |
| | | Vaccinated with 2nd dose, outside risk window | 28,583 | 16 | 114.43 | 1.01 | 0.59–1.72 | 0.975 | 0.92 | 0.53–1.58 | 0.751 |
| Facial nerve palsy | 28 days | *Overall* | 253,469 | 37 | 13.73 | | | | | | |
| | | Unvaccinated (ref.) | 252,759 | 18 | 21.96 | 1 | | | 1 | | |
| | | Vaccinated with 1st dose, inside risk window | 189,480 | < 5 | 13.77 | 0.63 | 0.15–2.70 | 0.532 | 0.76 | 0.16–3.57 | 0.731 |
| | | Vaccinated with 1st dose, outside risk window | 189,213 | 14 | 8.94 | 0.41 | 0.20–0.82 | 0.012 | 0.38 | 0.18–0.78 | 0.008 |
| | | Vaccinated with 2nd dose, inside risk window | 29,144 | 0 | 0.00 | ND | ND | ND | ND | ND | ND |
| | | Vaccinated with 2nd dose, outside risk window | 28,908 | < 5 | 21.19 | 0.96 | 0.28–3.28 | 0.954 | 0.73 | 0.21–2.59 | 0.626 |
| Guillain-Barré syndrome | 42 days | *Overall* | 253,663 | < 5 | 1.11 | | | | | | |
| | | Unvaccinated (ref.) | 252,953 | 0 | 0.00 | 1 | | | 1 | | |
| | | Vaccinated with 1st dose, inside risk window | 189,614 | 0 | 0.00 | ND | ND | ND | ND | ND | ND |
| | | Vaccinated with 1st dose, outside risk window | 189,144 | < 5 | 1.34 | ND | ND | ND | ND | ND | ND |
| | | Vaccinated with 2nd dose, inside risk window | 29,175 | 0 | 0.00 | ND | ND | ND | ND | ND | ND |
| | | Vaccinated with 2nd dose, outside risk window | 28,842 | < 5 | 7.65 | ND | ND | ND | ND | ND | ND |
| Henoch-Schönlein purpura | 42 days | *Overall* | 253,622 | < 10 | 2.22 | | | | | | |
| | | Unvaccinated (ref.) | 252,912 | < 5 | 1.22 | 1 | | | 1 | | |
| | | Vaccinated with 1st dose, inside risk window | 189,586 | 0 | 0.00 | ND | ND | ND | ND | ND | ND |
| | | Vaccinated with 1st dose, outside risk window | 189,116 | 5 | 3.35 | 2.74 | 0.32–23.49 | 0.357 | ND | ND | ND |
| | | Vaccinated with 2nd dose, inside risk window | 29,173 | 0 | 0.00 | ND | ND | ND | ND | ND | ND |
| | | Vaccinated with 2nd dose, outside risk window | 28,840 | 0 | 0.00 | ND | ND | ND | ND | ND | ND |
| Herpes zoster | 28 days | *Overall* | 253,556 | < 44 | 14.84 | | | | | | |
| | | Unvaccinated (ref.) | 252,846 | 10 | 12.19 | 1 | | | 1 | | |
| | | Vaccinated with 1st dose, inside risk window | 189,529 | < 5 | 6.88 | 0.56 | 0.07–4.41 | 0.586 | 1.23 | 0.13–11.20 | 0.857 |
| | | Vaccinated with 1st dose, outside risk window | 189,263 | 24 | 15.32 | 1.26 | 0.60–2.63 | 0.544 | 1.01 | 0.47–2.16 | 0.976 |
| | | Vaccinated with 2nd dose, inside risk window | 29,146 | 0 | 0.00 | ND | ND | ND | ND | ND | ND |
| | | Vaccinated with 2nd dose, outside risk window | 28,911 | 5 | 35.32 | 2.90 | 0.99–8.48 | 0.052 | 1.57 | 0.52–4.75 | 0.427 |
| Idiopathic thrombocytopenic purpura | 28 days | *Overall* | 253,601 | 13 | 4.82 | | | | | | |
| | | Unvaccinated (ref.) | 252,891 | < 5 | 3.66 | 1 | | | 1 | | |
| | | Vaccinated with 1st dose, inside risk window | 189,572 | < 5 | 13.77 | 3.76 | 0.63–22.51 | 0.147 | 3.06 | 0.42–22.09 | 0.267 |
| | | Vaccinated with 1st dose, outside risk window | 189,305 | 8 | 5.11 | 1.39 | 0.37–5.26 | 0.623 | 1.96 | 0.40–9.74 | 0.408 |
| | | Vaccinated with 2nd dose, inside risk window | 29,168 | 0 | 0.00 | ND | ND | ND | ND | ND | ND |
| | | Vaccinated with 2nd dose, outside risk window | 28,933 | 0 | 0.00 | ND | ND | ND | ND | ND | ND |
| Lymphadenopathy | 14 days | *Overall* | 252,508 | < 293 | 108.07 | | | | | | |
| | | Unvaccinated (ref.) | 251,803 | 74 | 90.65 | 1 | | | 1 | | |
| | | Vaccinated with 1st dose, inside risk window | 188,737 | 6 | 82.95 | 0.92 | 0.40–2.10 | 0.834 | 1.12 | 0.47–2.69 | 0.798 |
| | | Vaccinated with 1st dose, outside risk window | 188,706 | 185 | 113.37 | 1.25 | 0.96–1.64 | 0.104 | 1.15 | 0.87–1.52 | 0.338 |
| | | Vaccinated with 2nd dose, inside risk window | 28,993 | < 5 | 180.34 | 1.99 | 0.49–8.10 | 0.337 | 1.65 | 0.40–6.78 | 0.487 |
| | | Vaccinated with 2nd dose, outside risk window | 28,885 | 23 | 151.57 | 1.67 | 1.05–2.67 | 0.031 | 1.53 | 0.94–2.48 | 0.087 |
| Multisystem inflammatory syndrome in children | 42 days | *Overall* | 253,650 | 12 | 4.45 | | | | | | |
| | | Unvaccinated (ref.) | 252,940 | 5 | 6.09 | 1 | | | 1 | | |
| | | Vaccinated with 1st dose, inside risk window | 189,607 | 0 | 0.00 | ND | ND | ND | ND | ND | ND |
| | | Vaccinated with 1st dose, outside risk window | 189,137 | 7 | 4.68 | 0.77 | 0.24–2.42 | 0.652 | 1.16 | 0.29–4.55 | 0.834 |
| | | Vaccinated with 2nd dose, inside risk window | 29,176 | 0 | 0.00 | ND | ND | ND | ND | ND | ND |
| | | Vaccinated with 2nd dose, outside risk window | 28,843 | 0 | 0.00 | ND | ND | ND | ND | ND | ND |
| Myocarditis and pericarditis | 28 days | *Overall* | 253,659 | 10 | 3.71 | | | | | | |
| | | Unvaccinated (ref.) | 252,949 | < 5 | 3.66 | 1 | | | 1 | | |
| | | Vaccinated with 1st dose, inside risk window | 189,611 | 0 | 0.00 | ND | ND | ND | ND | ND | ND |
| | | Vaccinated with 1st dose, outside risk window | 189,346 | 5 | 3.19 | 0.87 | 0.21–3.65 | 0.852 | 1.30 | 0.25–6.75 | 0.757 |

| | | | | | | | | | | | |
|---|---|---|---|---|---|---|---|---|---|---|---|
| | | Vaccinated with 2nd dose, inside risk window | 29,174 | < 5 | 44.89 | 12.27 | 1.28–117.98 | 0.030 | 37.07 | 2.79–492.94 | 0.006 |
| | | Vaccinated with 2nd dose, outside risk window | 28,937 | < 5 | 7.06 | 1.93 | 0.20–18.54 | 0.569 | 4.12 | 0.31–54.93 | 0.285 |
| Venous thromboembolic events | 28 days | *Overall* | 253,650 | 16 | 5.93 | | | | | | |
| | | Unvaccinated (ref.) | 252,940 | < 5 | 2.44 | 1 | | | 1 | | |
| | | Vaccinated with 1st dose, inside risk window | 189,604 | 0 | 0.00 | ND | ND | ND | ND | ND | ND |
| | | Vaccinated with 1st dose, outside risk window | 189,339 | 13 | 8.29 | 3.41 | 0.77–15.10 | 0.107 | 6.03 | 0.78–46.37 | 0.084 |
| | | Vaccinated with 2nd dose, inside risk window | 29,169 | 0 | 0.00 | ND | ND | ND | ND | ND | ND |
| | | Vaccinated with 2nd dose, outside risk window | 28,933 | < 5 | 7.06 | 2.90 | 0.26–31.96 | 0.385 | 8.94 | 0.52–154.10 | 0.131 |

Abbreviations: CI – confidence interval; IRR – incidence rate ratio; mRNA – messenger RNA; ND – not determined

[1]12–15-year-olds: September 6, 2021; 16–17-year-olds: August 23, 2021; 18–19-year-olds: April 5, 2021

[2]Per 100,000 person-years

[3]Adjustment for sex (male or female), health region (North Norway, Central Norway, West Norway, or South-East Norway), and risk group (no or yes) as baseline covariates and three-month calendar period (April–June 2021, July–September 2021, October–December 2021, January–March 2022, April–June 2022, and July–September 2022) as a time-varying covariate

*Supplementary Table 3: Crude and adjusted incidence rate ratios of 17 different outcomes between vaccinated and unvaccinated subjects, with associated 95% confidence intervals, based on Poisson regression of 121,179 adolescents in Norway aged 16–17 years at the end of 2021 and unvaccinated against SARS-CoV-2 at the beginning of follow-up. Subjects were followed from the beginning of the wave of vaccination[1] of their age group until the outcome in question, non-mRNA SARS-CoV-2 vaccination, third-dose SARS-CoV-2 vaccination, emigration, death, or end of study on September 30, 2022, whichever occurred first. To ensure data privacy, numbers between 1 and 4 have been suppressed and are denoted by "< 5". As a result, some of the totals have been suppressed as well to avoid revealing small numbers that have been suppressed.*

| Outcome | Risk window | Vaccination status | Number of subjects | Number of events | Incidence rate[2] | Crude analysis | | | Adjusted analysis[3] | | |
|---|---|---|---|---|---|---|---|---|---|---|---|
| | | | | | | IRR | 95% CI | P value | IRR | 95% CI | P value |
| Acute appendicitis | 14 days | *Overall* | 120,117 | 300 | 227.97 | | | | | | |
| | | Unvaccinated (ref.) | 120,049 | 26 | 122.19 | 1 | | | 1 | | |
| | | Vaccinated with 1st dose, inside risk window | 106,881 | 7 | 170.88 | 1.40 | 0.61–3.22 | 0.431 | 1.07 | 0.45–2.56 | 0.878 |
| | | Vaccinated with 1st dose, outside risk window | 106,863 | 81 | 261.83 | 2.14 | 1.38–3.33 | 0.001 | 2.38 | 1.50–3.77 | 0.000 |
| | | Vaccinated with 2nd dose, inside risk window | 91,662 | 10 | 284.68 | 2.33 | 1.12–4.83 | 0.023 | 2.81 | 1.30–6.09 | 0.009 |
| | | Vaccinated with 2nd dose, outside risk window | 91,618 | 176 | 245.22 | 2.01 | 1.33–3.03 | 0.001 | 2.19 | 1.38–3.48 | 0.001 |
| Anaphylactic reaction | 2 days | *Overall* | 121,129 | < 17 | 10.54 | | | | | | |
| | | Unvaccinated (ref.) | 121,060 | < 5 | 9.32 | 1 | | | 1 | | |
| | | Vaccinated with 1st dose, inside risk window | 107,798 | 0 | 0.00 | ND | ND | ND | ND | ND | ND |
| | | Vaccinated with 1st dose, outside risk window | 107,796 | 5 | 14.37 | 1.54 | 0.30–7.96 | 0.604 | 1.79 | 0.34–9.43 | 0.495 |
| | | Vaccinated with 2nd dose, inside risk window | 92,475 | 0 | 0.00 | ND | ND | ND | ND | ND | ND |
| | | Vaccinated with 2nd dose, outside risk window | 92,474 | 7 | 9.27 | 0.99 | 0.21–4.79 | 0.995 | 0.82 | 0.15–4.55 | 0.823 |
| Arrhythmia | 28 days | *Overall* | 121,024 | < 36 | 24.11 | | | | | | |
| | | Unvaccinated (ref.) | 120,956 | 6 | 27.98 | 1 | | | 1 | | |
| | | Vaccinated with 1st dose, inside risk window | 107,700 | < 5 | 12.13 | 0.43 | 0.05–3.60 | 0.438 | 0.33 | 0.04–2.98 | 0.324 |
| | | Vaccinated with 1st dose, outside risk window | 106,724 | 6 | 22.15 | 0.79 | 0.26–2.45 | 0.685 | 1.23 | 0.32–4.66 | 0.763 |
| | | Vaccinated with 2nd dose, inside risk window | 92,396 | 0 | 0.00 | ND | ND | ND | ND | ND | ND |
| | | Vaccinated with 2nd dose, outside risk window | 92,318 | 19 | 27.59 | 0.99 | 0.39–2.47 | 0.976 | 1.63 | 0.47–5.64 | 0.437 |
| Arthropathy | 42 days | *Overall* | 121,179 | 0 | 0.00 | | | | | | |
| | | Unvaccinated (ref.) | 121,110 | 0 | 0.00 | 1 | | | 1 | | |
| | | Vaccinated with 1st dose, inside risk window | 107,846 | 0 | 0.00 | ND | ND | ND | ND | ND | ND |
| | | Vaccinated with 1st dose, outside risk window | 105,676 | 0 | 0.00 | ND | ND | ND | ND | ND | ND |
| | | Vaccinated with 2nd dose, inside risk window | 92,518 | 0 | 0.00 | ND | ND | ND | ND | ND | ND |
| | | Vaccinated with 2nd dose, outside risk window | 92,393 | 0 | 0.00 | ND | ND | ND | ND | ND | ND |
| Cerebrovascular events | 28 days | *Overall* | 121,144 | 11 | 8.28 | | | | | | |
| | | Unvaccinated (ref.) | 121,075 | < 5 | 9.32 | 1 | | | 1 | | |
| | | Vaccinated with 1st dose, inside risk window | 107,818 | < 5 | 36.34 | 3.90 | 0.65–23.33 | 0.136 | 5.52 | 0.71–42.97 | 0.102 |
| | | Vaccinated with 1st dose, outside risk window | 106,839 | < 5 | 7.37 | 0.79 | 0.11–5.61 | 0.815 | 0.61 | 0.08–4.51 | 0.630 |
| | | Vaccinated with 2nd dose, inside risk window | 92,488 | 0 | 0.00 | ND | ND | ND | ND | ND | ND |
| | | Vaccinated with 2nd dose, outside risk window | 92,410 | < 5 | 5.80 | 0.62 | 0.11–3.40 | 0.584 | 0.56 | 0.09–3.56 | 0.542 |
| Death | 28 days | *Overall* | 121,179 | < 31 | 21.82 | | | | | | |
| | | Unvaccinated (ref.) | 121,110 | 5 | 23.29 | 1 | | | 1 | | |
| | | Vaccinated with 1st dose, inside risk window | 107,846 | 0 | 0.00 | ND | ND | ND | ND | ND | ND |
| | | Vaccinated with 1st dose, outside risk window | 106,869 | 8 | 29.48 | 1.27 | 0.41–3.87 | 0.679 | 1.20 | 0.33–4.31 | 0.778 |
| | | Vaccinated with 2nd dose, inside risk window | 92,518 | < 5 | 42.31 | 1.82 | 0.43–7.60 | 0.414 | 1.42 | 0.28–7.17 | 0.668 |
| | | Vaccinated with 2nd dose, outside risk window | 92,440 | 13 | 18.85 | 0.81 | 0.29–2.27 | 0.687 | 1.16 | 0.32–4.23 | 0.818 |
| Encephalomyelitis and meningitis | 28 days | *Overall* | 121,166 | < 5 | 1.50 | | | | | | |
| | | Unvaccinated (ref.) | 121,097 | < 5 | 4.66 | 1 | | | 1 | | |
| | | Vaccinated with 1st dose, inside risk window | 107,834 | 0 | 0.00 | ND | ND | ND | ND | ND | ND |
| | | Vaccinated with 1st dose, outside risk window | 106,858 | < 5 | 3.69 | 0.79 | 0.05–12.64 | 0.868 | 0.60 | 0.04–10.25 | 0.726 |
| | | Vaccinated with 2nd dose, inside risk window | 92,508 | 0 | 0.00 | ND | ND | ND | ND | ND | ND |
| | | Vaccinated with 2nd dose, outside risk window | 92,431 | 0 | 0.00 | ND | ND | ND | ND | ND | ND |
| Epilepsy and convulsions | 28 days | *Overall* | 119,958 | 165 | 125.48 | | | | | | |

| | | | | | | | | | | | |
|---|---|---|---|---|---|---|---|---|---|---|---|
| | | Unvaccinated (ref.) | 119,891 | 26 | 122.46 | 1 | | | 1 | | |
| | | Vaccinated with 1st dose, inside risk window | 106,752 | 6 | 73.40 | 0.60 | 0.25–1.46 | 0.258 | 0.62 | 0.24–1.61 | 0.326 |
| | | Vaccinated with 1st dose, outside risk window | 105,783 | 34 | 126.69 | 1.03 | 0.62–1.72 | 0.896 | 0.92 | 0.53–1.60 | 0.766 |
| | | Vaccinated with 2nd dose, inside risk window | 91,573 | 15 | 213.77 | 1.75 | 0.92–3.30 | 0.086 | 1.36 | 0.68–2.73 | 0.383 |
| | | Vaccinated with 2nd dose, outside risk window | 91,482 | 84 | 123.11 | 1.01 | 0.65–1.56 | 0.981 | 1.00 | 0.60–1.67 | 0.997 |
| Facial nerve palsy | 28 days | *Overall* | 121,078 | 28 | 21.09 | | | | | | |
| | | Unvaccinated (ref.) | 121,009 | 5 | 23.31 | 1 | | | 1 | | |
| | | Vaccinated with 1st dose, inside risk window | 107,751 | < 5 | 12.12 | 0.52 | 0.06–4.45 | 0.550 | 0.26 | 0.03–2.29 | 0.225 |
| | | Vaccinated with 1st dose, outside risk window | 106,775 | 6 | 22.14 | 0.95 | 0.29–3.11 | 0.932 | 1.86 | 0.39–8.79 | 0.432 |
| | | Vaccinated with 2nd dose, inside risk window | 92,437 | < 5 | 28.23 | 1.21 | 0.23–6.24 | 0.819 | 2.07 | 0.28–15.52 | 0.480 |
| | | Vaccinated with 2nd dose, outside risk window | 92,357 | 14 | 20.32 | 0.87 | 0.31–2.42 | 0.792 | 3.15 | 0.62–16.12 | 0.168 |
| Guillain-Barré syndrome | 42 days | *Overall* | 121,172 | < 5 | 1.50 | | | | | | |
| | | Unvaccinated (ref.) | 121,103 | 0 | 0.00 | 1 | | | 1 | | |
| | | Vaccinated with 1st dose, inside risk window | 107,839 | 0 | 0.00 | ND | ND | ND | ND | ND | ND |
| | | Vaccinated with 1st dose, outside risk window | 105,669 | 0 | 0.00 | ND | ND | ND | ND | ND | ND |
| | | Vaccinated with 2nd dose, inside risk window | 92,512 | 0 | 0.00 | ND | ND | ND | ND | ND | ND |
| | | Vaccinated with 2nd dose, outside risk window | 92,387 | < 5 | 3.06 | ND | ND | ND | ND | ND | ND |
| Henoch-Schönlein purpura | 42 days | *Overall* | 121,166 | 6 | 4.51 | | | | | | |
| | | Unvaccinated (ref.) | 121,097 | < 5 | 4.66 | 1 | | | 1 | | |
| | | Vaccinated with 1st dose, inside risk window | 107,832 | 0 | 0.00 | ND | ND | ND | ND | ND | ND |
| | | Vaccinated with 1st dose, outside risk window | 105,662 | < 5 | 8.67 | 1.86 | 0.17–20.53 | 0.612 | 1.12 | 0.10–12.83 | 0.926 |
| | | Vaccinated with 2nd dose, inside risk window | 92,504 | 0 | 0.00 | ND | ND | ND | ND | ND | ND |
| | | Vaccinated with 2nd dose, outside risk window | 92,379 | < 5 | 4.59 | 0.98 | 0.10–9.46 | 0.989 | 0.78 | 0.06–9.44 | 0.843 |
| Herpes zoster | 28 days | *Overall* | 121,117 | 21 | 15.81 | | | | | | |
| | | Unvaccinated (ref.) | 121,048 | < 5 | 13.98 | 1 | | | 1 | | |
| | | Vaccinated with 1st dose, inside risk window | 107,792 | < 5 | 12.12 | 0.87 | 0.09–8.34 | 0.901 | 2.72 | 0.14–52.29 | 0.507 |
| | | Vaccinated with 1st dose, outside risk window | 106,815 | < 5 | 7.38 | 0.53 | 0.09–3.16 | 0.484 | 0.57 | 0.09–3.47 | 0.542 |
| | | Vaccinated with 2nd dose, inside risk window | 92,476 | 0 | 0.00 | ND | ND | ND | ND | ND | ND |
| | | Vaccinated with 2nd dose, outside risk window | 92,398 | 15 | 21.76 | 1.56 | 0.45–5.38 | 0.484 | 0.68 | 0.19–2.41 | 0.545 |
| Idiopathic thrombocytopenic purpura | 28 days | *Overall* | 121,150 | < 5 | 2.26 | | | | | | |
| | | Unvaccinated (ref.) | 121,081 | 0 | 0.00 | 1 | | | 1 | | |
| | | Vaccinated with 1st dose, inside risk window | 107,824 | 0 | 0.00 | ND | ND | ND | ND | ND | ND |
| | | Vaccinated with 1st dose, outside risk window | 106,848 | 0 | 0.00 | ND | ND | ND | ND | ND | ND |
| | | Vaccinated with 2nd dose, inside risk window | 92,501 | < 5 | 14.11 | ND | ND | ND | ND | ND | ND |
| | | Vaccinated with 2nd dose, outside risk window | 92,422 | < 5 | 2.90 | ND | ND | ND | ND | ND | ND |
| Lymphadenopathy | 14 days | *Overall* | 120,635 | < 181 | 135.37 | | | | | | |
| | | Unvaccinated (ref.) | 120,566 | 26 | 121.68 | 1 | | | 1 | | |
| | | Vaccinated with 1st dose, inside risk window | 107,344 | < 5 | 72.92 | 0.60 | 0.18–1.98 | 0.401 | 0.70 | 0.20–2.50 | 0.587 |
| | | Vaccinated with 1st dose, outside risk window | 107,330 | 49 | 157.61 | 1.30 | 0.81–2.08 | 0.286 | 1.16 | 0.71–1.91 | 0.548 |
| | | Vaccinated with 2nd dose, inside risk window | 92,070 | 10 | 283.42 | 2.33 | 1.12–4.83 | 0.023 | 1.90 | 0.87–4.13 | 0.105 |
| | | Vaccinated with 2nd dose, outside risk window | 92,026 | 91 | 126.16 | 1.04 | 0.67–1.60 | 0.871 | 0.95 | 0.58–1.54 | 0.825 |
| Multisystem inflammatory syndrome in children | 42 days | *Overall* | 121,176 | 8 | 6.02 | | | | | | |
| | | Unvaccinated (ref.) | 121,107 | < 5 | 4.66 | 1 | | | 1 | | |
| | | Vaccinated with 1st dose, inside risk window | 107,843 | < 5 | 24.33 | 5.22 | 0.54–50.23 | 0.152 | 5.99 | 0.48–75.25 | 0.166 |
| | | Vaccinated with 1st dose, outside risk window | 105,670 | < 5 | 4.34 | 0.93 | 0.06–14.89 | 0.960 | 0.74 | 0.05–12.22 | 0.836 |
| | | Vaccinated with 2nd dose, inside risk window | 92,516 | 0 | 0.00 | ND | ND | ND | ND | ND | ND |
| | | Vaccinated with 2nd dose, outside risk window | 92,391 | < 5 | 4.58 | 0.98 | 0.10–9.47 | 0.989 | 0.95 | 0.08–12.00 | 0.968 |
| Myocarditis and pericarditis | 28 days | *Overall* | 121,169 | 18 | 13.54 | | | | | | |
| | | Unvaccinated (ref.) | 121,100 | < 5 | 9.32 | 1 | | | 1 | | |
| | | Vaccinated with 1st dose, inside risk window | 107,839 | < 5 | 12.11 | 1.30 | 0.12–14.33 | 0.831 | 1.54 | 0.11–20.77 | 0.746 |
| | | Vaccinated with 1st dose, outside risk window | 106,861 | 7 | 25.80 | 2.77 | 0.58–13.33 | 0.204 | 2.15 | 0.41–11.17 | 0.362 |

| | | | | | | | | | | | |
|---|---|---|---|---|---|---|---|---|---|---|---|
| | | Vaccinated with 2nd dose, inside risk window | 92,509 | < 5 | 14.11 | 1.51 | 0.14–16.69 | 0.735 | 1.00 | 0.08–12.12 | 1.000 |
| | | Vaccinated with 2nd dose, outside risk window | 92,430 | 7 | 10.15 | 1.09 | 0.23–5.24 | 0.915 | 1.43 | 0.25–8.33 | 0.688 |
| Venous thromboembolic events | 28 days | *Overall* | 121,147 | 27 | 20.32 | | | | | | |
| | | Unvaccinated (ref.) | 121,078 | < 5 | 13.98 | 1 | | | 1 | | |
| | | Vaccinated with 1st dose, inside risk window | 107,816 | < 5 | 24.23 | 1.73 | 0.29–10.37 | 0.547 | 1.39 | 0.19–10.44 | 0.748 |
| | | Vaccinated with 1st dose, outside risk window | 106,837 | 5 | 18.43 | 1.32 | 0.32–5.52 | 0.705 | 1.51 | 0.31–7.41 | 0.609 |
| | | Vaccinated with 2nd dose, inside risk window | 92,488 | < 5 | 14.11 | 1.01 | 0.10–9.70 | 0.994 | 1.31 | 0.11–15.54 | 0.829 |
| | | Vaccinated with 2nd dose, outside risk window | 92,409 | 16 | 23.21 | 1.66 | 0.48–5.70 | 0.421 | 1.36 | 0.31–5.94 | 0.679 |

Abbreviations: CI – confidence interval; IRR – incidence rate ratio; mRNA – messenger RNA; ND – not determined

[1]12–15-year-olds: September 6, 2021; 16–17-year-olds: August 23, 2021; 18–19-year-olds: April 5, 2021

[2]Per 100,000 person-years

[3]Adjustment for sex (male or female), health region (North Norway, Central Norway, West Norway, or South-East Norway), and risk group (no or yes) as baseline covariates and three-month calendar period (April–June 2021, July–September 2021, October–December 2021, January–March 2022, April–June 2022, and July–September 2022) as a time-varying covariate

*Supplementary Table 4: Crude and adjusted incidence rate ratios of 17 different outcomes between vaccinated and unvaccinated subjects, with associated 95% confidence intervals, based on Poisson regression of 121,584 adolescents in Norway aged 18–19 years at the end of 2021 and unvaccinated against SARS-CoV-2 at the beginning of follow-up. Subjects were followed from the beginning of the wave of vaccination[1] of their age group until the outcome in question, non-mRNA SARS-CoV-2 vaccination, third-dose SARS-CoV-2 vaccination, emigration, death, or end of study on September 30, 2022, whichever occurred first. To ensure data privacy, numbers between 1 and 4 have been suppressed and are denoted by "< 5". As a result, some of the totals have been suppressed as well to avoid revealing small numbers that have been suppressed.*

| Outcome | Risk window | Vaccination status | Number of subjects | Number of events | Incidence rate[2] | Crude analysis | | | Adjusted analysis[3] | | |
|---|---|---|---|---|---|---|---|---|---|---|---|
| | | | | | | IRR | 95% CI | P value | IRR | 95% CI | P value |
| Acute appendicitis | 14 days | *Overall* | 120,562 | 349 | 250.73 | | | | | | |
| | | Unvaccinated (ref.) | 120,558 | 118 | 267.23 | 1 | | | 1 | | |
| | | Vaccinated with $1^{st}$ dose, inside risk window | 110,711 | 11 | 259.25 | 0.97 | 0.52–1.80 | 0.923 | 0.94 | 0.49–1.81 | 0.859 |
| | | Vaccinated with $1^{st}$ dose, outside risk window | 110,671 | 47 | 273.35 | 1.02 | 0.73–1.43 | 0.895 | 1.09 | 0.72–1.64 | 0.696 |
| | | Vaccinated with $2^{nd}$ dose, inside risk window | 104,941 | 9 | 223.78 | 0.84 | 0.43–1.65 | 0.608 | 0.83 | 0.40–1.71 | 0.607 |
| | | Vaccinated with $2^{nd}$ dose, outside risk window | 104,910 | 164 | 235.70 | 0.88 | 0.70–1.12 | 0.298 | 1.02 | 0.71–1.46 | 0.929 |
| Anaphylactic reaction | 2 days | *Overall* | 121,518 | < 25 | 14.94 | | | | | | |
| | | Unvaccinated (ref.) | 121,514 | 5 | 11.23 | 1 | | | 1 | | |
| | | Vaccinated with $1^{st}$ dose, inside risk window | 111,693 | 0 | 0.00 | ND | ND | ND | ND | ND | ND |
| | | Vaccinated with $1^{st}$ dose, outside risk window | 111,690 | 8 | 38.05 | 3.39 | 1.11–10.36 | 0.032 | 8.49 | 1.91–37.66 | 0.005 |
| | | Vaccinated with $2^{nd}$ dose, inside risk window | 105,912 | < 5 | 172.43 | 15.36 | 1.79–131.46 | 0.013 | 38.78 | 3.46–434.39 | 0.003 |
| | | Vaccinated with $2^{nd}$ dose, outside risk window | 105,909 | 7 | 9.49 | 0.84 | 0.27–2.66 | 0.774 | 2.10 | 0.37–12.07 | 0.406 |
| Arrhythmia | 28 days | *Overall* | 121,337 | < 57 | 38.49 | | | | | | |
| | | Unvaccinated (ref.) | 121,333 | 19 | 42.73 | 1 | | | 1 | | |
| | | Vaccinated with $1^{st}$ dose, inside risk window | 111,517 | < 5 | 23.55 | 0.55 | 0.13–2.37 | 0.422 | 0.52 | 0.11–2.50 | 0.418 |
| | | Vaccinated with $1^{st}$ dose, outside risk window | 101,548 | 8 | 61.02 | 1.43 | 0.63–3.26 | 0.398 | 1.06 | 0.42–2.66 | 0.907 |
| | | Vaccinated with $2^{nd}$ dose, inside risk window | 105,749 | 0 | 0.00 | ND | ND | ND | ND | ND | ND |
| | | Vaccinated with $2^{nd}$ dose, outside risk window | 105,684 | 25 | 37.80 | 0.88 | 0.49–1.61 | 0.687 | 0.58 | 0.27–1.26 | 0.169 |
| Arthropathy | 42 days | *Overall* | 121,569 | 0 | 0.00 | | | | | | |
| | | Unvaccinated (ref.) | 121,565 | 0 | 0.00 | 1 | | | 1 | | |
| | | Vaccinated with $1^{st}$ dose, inside risk window | 111,743 | 0 | 0.00 | ND | ND | ND | ND | ND | ND |
| | | Vaccinated with $1^{st}$ dose, outside risk window | 67,275 | 0 | 0.00 | ND | ND | ND | ND | ND | ND |
| | | Vaccinated with $2^{nd}$ dose, inside risk window | 105,968 | 0 | 0.00 | ND | ND | ND | ND | ND | ND |
| | | Vaccinated with $2^{nd}$ dose, outside risk window | 105,857 | 0 | 0.00 | ND | ND | ND | ND | ND | ND |
| Cerebrovascular events | 28 days | *Overall* | 121,533 | 6 | 4.27 | | | | | | |
| | | Unvaccinated (ref.) | 121,529 | < 5 | 8.98 | 1 | | | 1 | | |
| | | Vaccinated with $1^{st}$ dose, inside risk window | 111,706 | 0 | 0.00 | ND | ND | ND | ND | ND | ND |
| | | Vaccinated with $1^{st}$ dose, outside risk window | 101,722 | 0 | 0.00 | ND | ND | ND | ND | ND | ND |
| | | Vaccinated with $2^{nd}$ dose, inside risk window | 105,931 | 0 | 0.00 | ND | ND | ND | ND | ND | ND |
| | | Vaccinated with $2^{nd}$ dose, outside risk window | 105,866 | < 5 | 3.02 | 0.34 | 0.06–1.84 | 0.208 | 1.06 | 0.06–19.60 | 0.971 |
| Death | 28 days | *Overall* | 121,569 | 48 | 34.14 | | | | | | |
| | | Unvaccinated (ref.) | 121,565 | 18 | 40.41 | 1 | | | 1 | | |
| | | Vaccinated with $1^{st}$ dose, inside risk window | 111,743 | 6 | 70.52 | 1.75 | 0.69–4.40 | 0.237 | 1.53 | 0.52–4.50 | 0.439 |
| | | Vaccinated with $1^{st}$ dose, outside risk window | 101,754 | < 5 | 22.84 | 0.57 | 0.17–1.92 | 0.360 | 0.70 | 0.18–2.73 | 0.610 |
| | | Vaccinated with $2^{nd}$ dose, inside risk window | 105,968 | < 5 | 24.63 | 0.61 | 0.14–2.63 | 0.507 | 0.59 | 0.12–2.92 | 0.521 |
| | | Vaccinated with $2^{nd}$ dose, outside risk window | 105,903 | 19 | 28.67 | 0.71 | 0.37–1.35 | 0.297 | 1.30 | 0.46–3.66 | 0.623 |
| Encephalomyelitis and meningitis | 28 days | *Overall* | 121,551 | 7 | 4.98 | | | | | | |
| | | Unvaccinated (ref.) | 121,547 | < 5 | 2.25 | 1 | | | 1 | | |
| | | Vaccinated with $1^{st}$ dose, inside risk window | 111,727 | 0 | 0.00 | ND | ND | ND | ND | ND | ND |
| | | Vaccinated with $1^{st}$ dose, outside risk window | 101,739 | < 5 | 7.61 | 3.39 | 0.21–54.23 | 0.388 | 1.43 | 0.09–22.99 | 0.799 |
| | | Vaccinated with $2^{nd}$ dose, inside risk window | 105,952 | < 5 | 12.32 | 5.48 | 0.34–87.69 | 0.229 | 4.67 | 0.22–100.73 | 0.325 |

| | | | | | | | | | | | |
|---|---|---|---|---|---|---|---|---|---|---|---|
| | | Vaccinated with 2nd dose, outside risk window | 105,886 | < 5 | 6.04 | 2.69 | 0.30–24.06 | 0.376 | 0.80 | 0.09–7.48 | 0.846 |
| Epilepsy and convulsions | 28 days | *Overall* | 120,364 | 155 | 111.40 | | | | | | |
| | | Unvaccinated (ref.) | 120,360 | 47 | 106.53 | 1 | | | 1 | | |
| | | Vaccinated with 1st dose, inside risk window | 110,582 | 5 | 59.38 | 0.56 | 0.22–1.40 | 0.214 | 0.73 | 0.27–1.98 | 0.538 |
| | | Vaccinated with 1st dose, outside risk window | 100,711 | 21 | 161.49 | 1.52 | 0.91–2.54 | 0.113 | 1.85 | 0.97–3.54 | 0.064 |
| | | Vaccinated with 2nd dose, inside risk window | 104,852 | 14 | 174.23 | 1.64 | 0.90–2.97 | 0.106 | 2.20 | 1.05–4.61 | 0.036 |
| | | Vaccinated with 2nd dose, outside risk window | 104,776 | 68 | 103.72 | 0.97 | 0.67–1.41 | 0.888 | 1.08 | 0.60–1.93 | 0.806 |
| Facial nerve palsy | 28 days | *Overall* | 121,480 | < 30 | 18.51 | | | | | | |
| | | Unvaccinated (ref.) | 121,476 | 11 | 24.71 | 1 | | | 1 | | |
| | | Vaccinated with 1st dose, inside risk window | 111,654 | < 5 | 11.76 | 0.48 | 0.06–3.69 | 0.478 | 0.70 | 0.07–6.96 | 0.762 |
| | | Vaccinated with 1st dose, outside risk window | 101,671 | 0 | 0.00 | ND | ND | ND | ND | ND | ND |
| | | Vaccinated with 2nd dose, inside risk window | 105,888 | 0 | 0.00 | ND | ND | ND | ND | ND | ND |
| | | Vaccinated with 2nd dose, outside risk window | 105,823 | 14 | 21.14 | 0.86 | 0.39–1.88 | 0.699 | 0.48 | 0.16–1.40 | 0.180 |
| Guillain-Barré syndrome | 42 days | *Overall* | 121,566 | < 5 | 0.71 | | | | | | |
| | | Unvaccinated (ref.) | 121,562 | 0 | 0.00 | 1 | | | 1 | | |
| | | Vaccinated with 1st dose, inside risk window | 111,740 | 0 | 0.00 | ND | ND | ND | ND | ND | ND |
| | | Vaccinated with 1st dose, outside risk window | 67,273 | 0 | 0.00 | ND | ND | ND | ND | ND | ND |
| | | Vaccinated with 2nd dose, inside risk window | 105,965 | 0 | 0.00 | ND | ND | ND | ND | ND | ND |
| | | Vaccinated with 2nd dose, outside risk window | 105,854 | < 5 | 1.61 | ND | ND | ND | ND | ND | ND |
| Henoch-Schönlein purpura | 42 days | *Overall* | 121,555 | < 5 | 1.42 | | | | | | |
| | | Unvaccinated (ref.) | 121,551 | 0 | 0.00 | 1 | | | 1 | | |
| | | Vaccinated with 1st dose, inside risk window | 111,731 | 0 | 0.00 | ND | ND | ND | ND | ND | ND |
| | | Vaccinated with 1st dose, outside risk window | 67,270 | 0 | 0.00 | ND | ND | ND | ND | ND | ND |
| | | Vaccinated with 2nd dose, inside risk window | 105,956 | 0 | 0.00 | ND | ND | ND | ND | ND | ND |
| | | Vaccinated with 2nd dose, outside risk window | 105,845 | < 5 | 3.21 | ND | ND | ND | ND | ND | ND |
| Herpes zoster | 28 days | *Overall* | 121,508 | 19 | 13.52 | | | | | | |
| | | Unvaccinated (ref.) | 121,504 | 5 | 11.23 | 1 | | | 1 | | |
| | | Vaccinated with 1st dose, inside risk window | 111,680 | 0 | 0.00 | ND | ND | ND | ND | ND | ND |
| | | Vaccinated with 1st dose, outside risk window | 101,697 | < 5 | 22.85 | 2.04 | 0.49–8.52 | 0.330 | 1.70 | 0.31–9.34 | 0.544 |
| | | Vaccinated with 2nd dose, inside risk window | 105,908 | < 5 | 12.32 | 1.10 | 0.13–9.40 | 0.932 | 1.09 | 0.10–12.23 | 0.942 |
| | | Vaccinated with 2nd dose, outside risk window | 105,842 | 10 | 15.10 | 1.35 | 0.46–3.94 | 0.588 | 0.88 | 0.19–3.98 | 0.868 |
| Idiopathic thrombocytopenic purpura | 28 days | *Overall* | 121,538 | < 10 | 5.69 | | | | | | |
| | | Unvaccinated (ref.) | 121,534 | < 5 | 6.74 | 1 | | | 1 | | |
| | | Vaccinated with 1st dose, inside risk window | 111,711 | 0 | 0.00 | ND | ND | ND | ND | ND | ND |
| | | Vaccinated with 1st dose, outside risk window | 101,728 | 0 | 0.00 | ND | ND | ND | ND | ND | ND |
| | | Vaccinated with 2nd dose, inside risk window | 105,936 | 0 | 0.00 | ND | ND | ND | ND | ND | ND |
| | | Vaccinated with 2nd dose, outside risk window | 105,872 | 5 | 7.55 | 1.12 | 0.27–4.69 | 0.877 | 2.45 | 0.39–15.45 | 0.340 |
| Lymphadenopathy | 14 days | *Overall* | 120,995 | 182 | 130.15 | | | | | | |
| | | Unvaccinated (ref.) | 120,991 | 52 | 117.29 | 1 | | | 1 | | |
| | | Vaccinated with 1st dose, inside risk window | 111,165 | 6 | 140.83 | 1.20 | 0.52–2.80 | 0.671 | 1.30 | 0.53–3.16 | 0.568 |
| | | Vaccinated with 1st dose, outside risk window | 111,130 | 15 | 86.87 | 0.74 | 0.42–1.32 | 0.306 | 0.84 | 0.43–1.64 | 0.610 |
| | | Vaccinated with 2nd dose, inside risk window | 105,405 | 10 | 247.56 | 2.11 | 1.07–4.15 | 0.031 | 2.37 | 1.09–5.15 | 0.030 |
| | | Vaccinated with 2nd dose, outside risk window | 105,373 | 99 | 141.57 | 1.21 | 0.86–1.69 | 0.272 | 1.30 | 0.77–2.20 | 0.330 |
| Multisystem inflammatory syndrome in children | 42 days | *Overall* | 121,563 | < 5 | 0.71 | | | | | | |
| | | Unvaccinated (ref.) | 121,559 | 0 | 0.00 | 1 | | | 1 | | |
| | | Vaccinated with 1st dose, inside risk window | 111,737 | 0 | 0.00 | ND | ND | ND | ND | ND | ND |
| | | Vaccinated with 1st dose, outside risk window | 67,272 | 0 | 0.00 | ND | ND | ND | ND | ND | ND |
| | | Vaccinated with 2nd dose, inside risk window | 105,964 | 0 | 0.00 | ND | ND | ND | ND | ND | ND |
| | | Vaccinated with 2nd dose, outside risk window | 105,853 | < 5 | 1.61 | ND | ND | ND | ND | ND | ND |
| Myocarditis and pericarditis | 28 days | *Overall* | 121,531 | < 41 | 27.04 | | | | | | |
| | | Unvaccinated (ref.) | 121,527 | 8 | 17.96 | 1 | | | 1 | | |

| | | | | | | | | | | | |
|---|---|---|---|---|---|---|---|---|---|---|---|
| | | Vaccinated with 1st dose, inside risk window | 111,700 | < 5 | 23.51 | 1.31 | 0.28–6.16 | 0.733 | 1.87 | 0.31–11.29 | 0.493 |
| | | Vaccinated with 1st dose, outside risk window | 101,714 | 5 | 38.10 | 2.12 | 0.69–6.48 | 0.187 | 3.91 | 0.81–18.77 | 0.089 |
| | | Vaccinated with 2nd dose, inside risk window | 105,926 | 9 | 110.87 | 6.17 | 2.38–16.00 | < 0.001 | 10.25 | 2.36–44.47 | 0.002 |
| | | Vaccinated with 2nd dose, outside risk window | 105,852 | 14 | 21.14 | 1.18 | 0.49–2.81 | 0.713 | 2.84 | 0.61–13.15 | 0.182 |
| Venous thromboembolic events | 28 days | *Overall* | 121,513 | 26 | 18.50 | | | | | | |
| | | Unvaccinated (ref.) | 121,509 | 6 | 13.47 | 1 | | | 1 | | |
| | | Vaccinated with 1st dose, inside risk window | 111,684 | < 5 | 35.28 | 2.62 | 0.65–10.47 | 0.173 | 5.07 | 0.96–26.81 | 0.056 |
| | | Vaccinated with 1st dose, outside risk window | 101,699 | < 5 | 15.23 | 1.13 | 0.23–5.60 | 0.881 | 2.39 | 0.31–18.51 | 0.403 |
| | | Vaccinated with 2nd dose, inside risk window | 105,907 | < 5 | 24.64 | 1.83 | 0.37–9.06 | 0.460 | 3.99 | 0.52–30.36 | 0.181 |
| | | Vaccinated with 2nd dose, outside risk window | 105,840 | 13 | 19.63 | 1.46 | 0.55–3.83 | 0.446 | 2.59 | 0.43–15.41 | 0.296 |

Abbreviations: CI – confidence interval; IRR – incidence rate ratio; mRNA – messenger RNA; ND – not determined

[1]12–15-year-olds: September 6, 2021; 16–17-year-olds: August 23, 2021; 18–19-year-olds: April 5, 2021

[2]Per 100,000 person-years

[3]Adjustment for sex (male or female), health region (North Norway, Central Norway, West Norway, or South-East Norway), and risk group (no or yes) as baseline covariates and three-month calendar period (April–June 2021, July–September 2021, October–December 2021, January–March 2022, April–June 2022, and July–September 2022) as a time-varying covariate

*Supplementary Table 5: Adjusted incidence rate ratios of 17 different outcomes between vaccinated and unvaccinated subjects, with associated 95% confidence intervals, based on self-controlled case series (SCCS) analysis of adolescents in Norway aged 12–19 years at the end of 2021 and unvaccinated against SARS-CoV-2 at the beginning of follow-up.*

| Outcome | Risk window | Vaccination status | Number of events | Adjusted analysis[1] | | |
|---|---|---|---|---|---|---|
| | | | | IRR | 95% CI | P value |
| Acute appendicitis | 14 days | *Total* | 1,185 | | | |
| | | Unvaccinated (ref.) | 267 | 1 | | |
| | | Vaccinated with 1st dose, inside risk window | 36 | 1.05 | 0.72–1.54 | 0.79 |
| | | Vaccinated with 1st dose, outside risk window | 495 | 1.15 | 0.90–1.45 | 0.26 |
| | | Vaccinated with 2nd dose, inside risk window | 21 | 1.02 | 0.64–1.63 | 0.94 |
| | | Vaccinated with 2nd dose, outside risk window | 366 | 0.97 | 0.78–1.22 | 0.81 |
| Anaphylactic reaction | 2 days | *Total* | < 61 | | | |
| | | Unvaccinated (ref.) | 12 | 1 | | |
| | | Vaccinated with 1st dose, inside risk window | < 5 | 4.92 | 0.59–41.11 | 0.14 |
| | | Vaccinated with 1st dose, outside risk window | 26 | 2.02 | 0.74–5.49 | 0.17 |
| | | Vaccinated with 2nd dose, inside risk window | < 5 | 2.02 | 0.89–64.17 | 0.06 |
| | | Vaccinated with 2nd dose, outside risk window | 16 | 0.77 | 0.29–2.03 | 0.60 |
| Arrhythmia | 28 days | *Total* | 135 | | | |
| | | Unvaccinated (ref.) | 39 | 1 | | |
| | | Vaccinated with 1st dose, inside risk window | < 5 | 0.48 | 0.16–1.41 | 0.18 |
| | | Vaccinated with 1st dose, outside risk window | 43 | 1.01 | 0.53–1.91 | 0.99 |
| | | Vaccinated with 2nd dose, inside risk window | < 5 | 0.19 | 0.03–1.45 | 0.11 |
| | | Vaccinated with 2nd dose, outside risk window | 48 | 1.30 | 0.71–2.39 | 0.39 |
| Arthropathy | 42 days | *Total* | 0 | | | |
| | | Unvaccinated (ref.) | 0 | 1 | | |
| | | Vaccinated with 1st dose, inside risk window | 0 | ND | ND | ND |
| | | Vaccinated with 1st dose, outside risk window | 0 | ND | ND | ND |
| | | Vaccinated with 2nd dose, inside risk window | 0 | ND | ND | ND |
| | | Vaccinated with 2nd dose, outside risk window | 0 | ND | ND | ND |
| Cerebrovascular events | 28 days | *Total* | < 33 | | | |
| | | Unvaccinated (ref.) | 10 | 1 | | |
| | | Vaccinated with 1st dose, inside risk window | < 5 | 1.22 | 0.26–5.72 | 0.80 |

| | | | | | | |
|---|---|---|---|---|---|---|
| | | Vaccinated with 1st dose, outside risk window | 12 | 0.80 | 0.22–2.92 | 0.74 |
| | | Vaccinated with 2nd dose, inside risk window | 0 | ND | ND | ND |
| | | Vaccinated with 2nd dose, outside risk window | 6 | 0.44 | 0.11–1.75 | 0.24 |
| Death | 28 days | *Total* | | | | |
| | | Unvaccinated (ref.) | | 1 | | |
| | | Vaccinated with 1st dose, inside risk window | | ND | ND | ND |
| | | Vaccinated with 1st dose, outside risk window | | ND | ND | ND |
| | | Vaccinated with 2nd dose, inside risk window | | ND | ND | ND |
| | | Vaccinated with 2nd dose, outside risk window | | ND | ND | ND |
| Encephalomyelitis and meningitis | 28 days | *Total* | 14 | | | |
| | | Unvaccinated (ref.) | < 5 | 1 | | |
| | | Vaccinated with 1st dose, inside risk window | 0 | ND | ND | ND |
| | | Vaccinated with 1st dose, outside risk window | 5 | ND | ND | ND |
| | | Vaccinated with 2nd dose, inside risk window | < 5 | ND | ND | ND |
| | | Vaccinated with 2nd dose, outside risk window | < 5 | ND | ND | ND |
| Epilepsy and convulsions | 28 days | *Total* | 617 | | | |
| | | Unvaccinated (ref.) | 165 | 1 | | |
| | | Vaccinated with 1st dose, inside risk window | 29 | 0.95 | 0.59–1.52 | 0.83 |
| | | Vaccinated with 1st dose, outside risk window | 225 | 1.08 | 0.76–1.56 | 0.66 |
| | | Vaccinated with 2nd dose, inside risk window | 30 | 1.59 | 0.99–2.55 | 0.05 |
| | | Vaccinated with 2nd dose, outside risk window | 168 | 1.08 | 0.77–1.52 | 0.66 |
| Facial nerve palsy | 28 days | *Total* | 91 | | | |
| | | Unvaccinated (ref.) | 34 | 1 | | |
| | | Vaccinated with 1st dose, inside risk window | < 5 | 0.89 | 0.27–2.96 | 0.85 |
| | | Vaccinated with 1st dose, outside risk window | 20 | 0.96 | 0.37–2.48 | 0.93 |
| | | Vaccinated with 2nd dose, inside risk window | < 5 | 0.64 | 0.13–3.04 | 0.57 |
| | | Vaccinated with 2nd dose, outside risk window | 31 | 1.11 | 0.49-2.52 | 0.81 |
| Guillain-Barré syndrome | 42 days | *Total* | 6 | | | |
| | | Unvaccinated (ref.) | 0 | 1 | | |
| | | Vaccinated with 1st dose, inside risk window | 0 | ND | ND | ND |
| | | Vaccinated with 1st dose, outside risk window | < 5 | ND | ND | ND |
| | | Vaccinated with 2nd dose, inside risk window | 0 | ND | ND | ND |
| | | Vaccinated with 2nd dose, outside risk window | < 5 | ND | ND | ND |

| | | | | | | |
|---|---|---|---|---|---|---|
| Henoch-Schönlein purpura | 42 days | *Total* | < 17 | | | |
| | | Unvaccinated (ref.) | < 5 | 1 | | |
| | | Vaccinated with 1st dose, inside risk window | 0 | ND | ND | ND |
| | | Vaccinated with 1st dose, outside risk window | 7 | 0.63 | 0.07–5.44 | 0.67 |
| | | Vaccinated with 2nd dose, inside risk window | 0 | ND | ND | ND |
| | | Vaccinated with 2nd dose, outside risk window | 5 | 1.48 | 0.12–18.91 | 0.76 |
| Herpes zoster | 28 days | *Total* | 80 | | | |
| | | Unvaccinated (ref.) | 18 | 1 | | |
| | | Vaccinated with 1st dose, inside risk window | < 5 | 1.56 | 0.25–9.67 | 0.63 |
| | | Vaccinated with 1st dose, outside risk window | 29 | 2.26 | 0.56–9.19 | 0.25 |
| | | Vaccinated with 2nd dose, inside risk window | < 5 | 1.65 | 0.16–16.80 | 0.67 |
| | | Vaccinated with 2nd dose, outside risk window | 30 | 4.12 | 1.10–15.4 | 0.04 |
| Idiopathic thrombocytopenic purpura | 28 days | *Total* | 24 | | | |
| | | Unvaccinated (ref.) | 6 | 1 | | |
| | | Vaccinated with 1st dose, inside risk window | < 5 | 0.75 | 0.18–10.66 | 0.75 |
| | | Vaccinated with 1st dose, outside risk window | 8 | 0.89 | 0.12–6.78 | 0.91 |
| | | Vaccinated with 2nd dose, inside risk window | < 5 | 1.21 | 0.10-13.89 | 0.88 |
| | | Vaccinated with 2nd dose, outside risk window | 7 | 1.09 | 0.19–6.41 | 0.92 |
| Lymphadenopathy | 14 days | *Total* | 651 | | | |
| | | Unvaccinated (ref.) | 152 | 1 | | |
| | | Vaccinated with 1st dose, inside risk window | 15 | 0.79 | 0.45–1.40 | 0.43 |
| | | Vaccinated with 1st dose, outside risk window | 249 | 1.00 | 0.73–1.38 | 0.98 |
| | | Vaccinated with 2nd dose, inside risk window | 22 | 2.04 | 1.24–3.35 | 0.01 |
| | | Vaccinated with 2nd dose, outside risk window | 213 | 1.08 | 0.80–1.47 | 0.61 |
| Multisystem inflammatory syndrome in children | 42 days | *Total* | 21 | | | |
| | | Unvaccinated (ref.) | 6 | 1 | | |
| | | Vaccinated with 1st dose, inside risk window | < 5 | 1.14 | 0.17–7.52 | 0.89 |
| | | Vaccinated with 1st dose, outside risk window | 8 | 0.47 | 0.09–2.62 | 0.39 |
| | | Vaccinated with 2nd dose, inside risk window | 0 | NA | NA | NA |
| | | Vaccinated with 2nd dose, outside risk window | < 5 | 1.33 | 0.11–16.65 | 0.82 |
| Myocarditis and pericarditis | 28 days | *Total* | < 68 | | | |
| | | Unvaccinated (ref.) | 13 | 1 | | |
| | | Vaccinated with 1st dose, inside risk window | < 5 | 1.20 | 0.30–4.84 | 0.80 |

| | | | | | | |
|---|---|---|---|---|---|---|
| | | Vaccinated with 1st dose, outside risk window | 17 | 1.60 | 0.53–4.84 | 0.40 |
| | | Vaccinated with 2nd dose, inside risk window | 11 | 5.88 | 2.11–16.40 | <0.001 |
| | | Vaccinated with 2nd dose, outside risk window | 22 | 1.14 | 0.45–2.93 | 0.78 |
| Venous thromboembolic events | 28 days | *Total* | < 71 | | | |
| | | Unvaccinated (ref.) | 11 | 1 | | |
| | | Vaccinated with 1st dose, inside risk window | 5 | 1.05 | 0.35–3.16 | 0.93 |
| | | Vaccinated with 1st dose, outside risk window | 20 | 1.04 | 0.42–2.62 | 0.93 |
| | | Vaccinated with 2nd dose, inside risk window | < 5 | 0.81 | 0.22–3.02 | 0.75 |
| | | Vaccinated with 2nd dose, outside risk window | 30 | 0.98 | 0.45–2.14 | 0.95 |

[1]Adjustment for seasonality, defined as quarterly time periods (January–March, April–June, July–September, and October–December)
